# Influence of Genetic Ancestry on Gene-Environment Interactions of Polygenic Risk and Sociocultural Factors: Results from the Hispanic Community Health Study/Study of Latinos

**DOI:** 10.1101/2024.11.26.24318009

**Authors:** Jayati Sharma, Cristin E. McArdle, Mariaelisa Graff, Christina Cordero, Martha Daviglus, Linda C. Gallo, Carmen R. Isasi, Tanika N. Kelly, Krista M. Perreira, Gregory A. Talavera, Jianwen Cai, Kari E. North, Lindsay Fernández-Rhodes, Genevieve L. Wojcik

## Abstract

**Background:** Many present analyses of Hispanic/Latino populations in epidemiologic research aggregate all members of this ethnic group, despite immense diversity in genetic backgrounds, environment, and culture between and across Hispanic/Latino background groups. Using the Hispanic Community Health Study/Study of Latinos (HCHS/SOL), we examined the role of self-identified background group and genetic ancestry proportions in gene-environment interactions influencing the relationship between body mass index (BMI) and a polygenic score for BMI (PGS_BMI_).

**Methods:** Weighted univariate and multivariable generalized linear models were executed to compare the effects of environmental variables identified *a priori* by McArdle et al. 2021. Both Amerindigenous (AME) ancestry proportion and background group identity were statistically modeled as confounders both through stratified and joint analyses to understand their influence on the relationship between BMI and PGS_BMI_, while incorporating gene-environment interactions of PGS x diet and PGS x age-at-immigration.

**Results:** After complex survey weighting, 7,075 participants remained in the analytic sample, representing individuals of six background groups: Central American, Cuban, Dominican, Mexican, Puerto Rican, and South American. The distributions of key environmental and sociocultural variables were heterogeneous between Hispanic/Latino background groups. Associations of these variables with AME ancestry were similarly heterogeneous upon stratification, indicating confounding by background group. In a predictive model for BMI incorporating health, immigration, and environmental variables, PGS_BMI_ performance decreased with increasing AME ancestry proportion. In this model, most statistically significant GxE interactions lost significance after ancestry and background stratification, except for PGS x age-at-immigration interactions in some strata: Mexican background individuals born in the US compared to those >=21 years old at migration (β=1.33, p<0.01), Dominican background individuals 6-12 years old at migration compared to those >=21 years old at migration (β=4.38, p<0.001), and Cuban background individuals 0-5 years old at migration compared to those >=21 years old at migration (β=2.20, p=0.015), where US-born includes individuals born in the US 50 states/DC.

**Conclusions:** Controlling for self-identified background group identity and genetic ancestry did not eliminate statistically significant differences in interactions between AME ancestry and environmental variables in certain strata of AME ancestry among some Hispanic/Latino background groups in HCHS/SOL.

## INTRODUCTION

Hispanic/Latino groups in the United States (US) comprise an inherently diverse ethnicity, and yet are often modeled as a monolithic group in epidemiologic analyses, despite their unique sociocultural and genetic composition. This issue is especially prevalent in presentation of race- and ethnicity-stratified health and socioeconomic statistics in the US, where all members of Hispanic/Latino ethnicity are aggregated into a singular category regardless of their specific Hispanic/Latino background or birthplace.^1,2^ In a systematic review and meta-analysis of cardiovascular mortality in Hispanic/Latino populations in the US that identified lower mortality among Hispanic/Latino individuals than their non-Hispanic white counterparts, authors identified only one of 17 included studies that stratified participants by place of birth.^3^ Recent literature examining the role of acculturation and heterogeneous sociocultural landscapes within Hispanic/Latino backgrounds has started to address this scarcity, through the study of topics ranging from substance-use treatment outcomes to food insecurity and obesity research.^4,5^ These important distinctions are also reflected in the genetic diversity within the Hispanic/Latino ethnicity, evidenced in various analyses of population structure and genetic ancestry proportions of Hispanic/Latino populations.^6,7^ Diverse Hispanic/Latino experiences and histories, including complex geographic histories shaped through colonization and immigration, have and continue to shape the cultures, behaviors, and health of Hispanic/Latino groups throughout the US.

As the second-largest ethnic or racial group in the US and as a historically marginalized population, Hispanic/Latino groups are the subject of many public health studies focused on health inequities.^8^ One facet of these inequities is in the burden of chronic diseases, exacerbated by obesity, which 44.8% of adult Hispanic/Latino individuals in the US face.^9^ While lifestyle factors and health behaviors have been studied as predictors of obesity, the incorporation of genetics and gene-environment interactions are another promising avenue through which to understand the individual-level impact of these factors.^10^ Gene-environment (GxE) interactions, which characterize the joint influence of genetic and environmental variables (such as health behaviors), are an important area of study that may inform future precision health applications of screenings and therapeutics designed to prevent and treat chronic diseases caused by pre-existing conditions such as obesity. In Hispanic/Latino populations, disproportionate levels of exposure to obesogenic environments via poor diet quality, low socioeconomic status, poor education, and healthcare bias may interact with obesity-associated genetic polymorphisms and may contribute to group-level disparities by Hispanic/Latino background.^11^

Studying disease disparities and GxE interactions across populations, particularly of diverse genetic ancestries, can advance our understanding of the complex relationships between genetic and lifestyle or behavioral factors that may have been otherwise unobservable in non-genetic analyses. One approach to examining genetic ancestry is by estimating admixture proportions, in which each individual’s genome is apportioned based on its similarity to either a reference population, or other genomic segments in the sample.^12,13^ Proportions of inferred genetic ancestry, or admixture proportions, have been shown to vary widely by Hispanic/Latino background, typically modeled with European, African, and Native American (AME; referred to in this manuscript as Amerindigenous) ancestry components showing higher African ancestry in Caribbean populations (Dominican, Puerto Rican) and higher Amerindigenous ancestry in Mexican participants.^14,15^ Reflecting differences in migration patterns, differences in ancestry proportions have also been seen to persist based on geographic position in the United States when examined across Hispanic/Latino participants.^16^

Recently, an effective way to model the genetic liability of complex traits and diseases is through polygenic scores (PGS), a relatively comprehensive metric comprising a weighted sum of many genetic variants (single nucleotide polymorphisms, or SNPs) associated with a given particular trait. However, there are noted challenges with the use of PGS across diverse populations. Recent research has identified the decreased PGS_BMI_ performance in non-European participants, in particular those from African, South Asian, East Asian, and Hispanic/Latino backgrounds, when samples of European ancestries are used to train such PGSs.^17^ This may be due to differences in the underlying genetic architecture, including linkage disequilibrium patterns and allele frequencies, but could also be due to differing environmental influences.^18^ Decreased PGS performance has also been observed when examining non-genetic factors that differ between training and test sets, such as age and sex.^19,20^ However, there is an open question as to the degree to which ancestry and environmental differences jointly contribute to PGS performance.

The Hispanic Community Health Study/Study of Latinos (HCHS/SOL) is an ideal cohort to examine these relationships as participants provide background information during study enrollment as well as extensive well-characterized longitudinal data. A previous analysis by McArdle *et al.* examined the influence of PGS-by-diet and PGS-by-acculturation interactions on the genetic risk of obesity via a PGS_BMI_ trained on European ancestry samples from UK Biobank and GIANT and applied in HCHS/SOL.^21^ The authors found dietary patterns and age at immigration to be significant modifiers of the effect of individuals’ genetic risk on obesity; the effect of the PGS_BMI_ on BMI was different based on different levels of acculturation and healthy diet. Specifically, in their full model, the authors identified that a one-standard deviation (SD) increase in the PGS_BMI_ was associated with a 1.10 kg/m^2^ increase in BMI (β=1.10,p<0.001), adjusted for various demographic, sociocultural, and environmental variables, which differed substantially when stratified by sex (males: β=0.79, p<0.001; females: β=1.45, p<0.001). A separate exploratory analysis stratified by self-reported Hispanic/Latino background observed significant heterogeneity for PGS_BMI_, with weaker effects in individuals of South American background (β=0.91, p<0.001) than of Mexican background (β=1.73, p<0.001). Complicating this, the modifying effects of age-at-immigration and healthy diet on PGS_BMI_ showed different direction of effects across background groups, such as when comparing individuals were born in the US 50 states/DC to those having migrated to the US after the age of 20 years old (South American background: β=-1.74,p<0.05; Mexican background: β=1.09,p<0.05).

It remains unclear, however, the extent to which background within Hispanic/Latino ethnicity influences these relationships and what factors would influence these interactions. Given this discrepancy in predictive performance, we hypothesize that there exist ancestry-driven differences in the performance of this PGS_BMI_ in Hispanic/Latino populations. In addition, it is unclear how the intersection of ancestry and environmental differences may influence the performance of a PGS. Building on prior work, we hypothesize that the GxE interactions between a PGS_BMI_ and immigration history and diet variables varied between background groups of Hispanic/Latino ethnicity as a function of both group-specific ancestry differences as well as differences in environment. There is an urgent need to disentangle these complex factors to better characterize the joint roles of genetics and environmental influences on human health, as well as demonstrate the need to model Hispanic/Latino populations appropriately in genetic research. As such, this paper broadly seeks to (1) better understand the role of inferred genetic ancestry with BMI, both between and within Hispanic/Latino groups, and (2) expand the analysis of McArdle *et al*. to incorporate AME ancestry proportion and examine the influence of group heterogeneity on interactions between the afore-mentioned sociocultural variables and genetic risk for obesity.

## METHODS

### Study Design

The HCHS/SOL study design and methodology have been described in detail elsewhere.^22^ From four sites (Bronx, Miami, Chicago, San Diego) in the US, 16,415 self-identified Hispanic/Latino participants aged 18-74 were recruited and had physical, behavioral, sociocultural, and biometric measurements collected at a baseline examination between 2008-2011 and second clinic visit between 2014-2017. Participants were recruited to HCHS/SOL through a two-stage area household probability design and therefore some participants are related.^22^

In the present study, we examined data from an analytic subset of HCHS/SOL participants who were included in the analysis conducted by McArdle et al.^21^: those who had consented for genetic data collection (at visit 1) and analysis (at both visits), and whose information was successfully linked to the PGS_BMI_ data constructed in HCHS/SOL. This analytic subset was then further restricted to participants with visit 1 data, with estimated admixture proportions for genetic ancestry, and no missing covariate data (n=7,282). The inclusion/exclusion criteria and the resulting sample size of this study are described in Supplementary Figure 1.

### Variable Definitions

Immigration-related variables used in this analysis were immigrant generation (first or second) and age at immigration to the US. Immigrant generation was defined as "first generation" as being foreign-born with foreign-born parents, including those born in a US territory. "Second generation" was defined as those who were born in the US or those who were foreign-born with at least one US-born parent. Age at immigration to the US was defined among the foreign/territory-born participants based on their current age and years residing in the US. Following McArdle *et al*., five categories were defined: Born in US 50 states/DC (hereafter, US born), 0-5 years old at migration, 6-12 years old at migration, 13-20 years old at migration, and ≥21 years old at migration.

Several other health and environmental variables were also included as covariates in the full model, derived from the analysis conducted by McArdle et al. These consisted of sex (binary: male or female); age; age^2^; self-reported Hispanic/Latino background identity and descent (Central American, Cuban, Dominican, Mexican, Puerto Rican, and South American); marital status (currently married, yes/no); educational status (less than high school vs. high school and beyond); employment status (retired, not retired or employed, employed ≤ 35 h/week, employed > 35 h/week); type 2 diabetes status (yes/no); cardiovascular disease (CVD) status, e.g. CHD or Stroke, but not counting angina or transient ischemic attacks (yes/no); sleep duration (h/day); sweetened beverages consumption (servings/day); whether the WHO’s 2008 Global Physical Activity guidelines for Americans criteria^23^ were met (yes/no); alcohol use level (no current use, low-level use, and high-level use); cigarette use (never, current, and former); Center for Epidemiological Studies Depression (CES-D) 10-item Summary Score^24^; and a single item ethnic identity score (5-level ordinal variable).

Healthy diet was defined using in part the JAMA Healthy Diet Score, which used data from the baseline 24-hour dietary recall to assign individuals a value ranging from 1 to 5 based on meeting sex-specific quintiles of predicted daily intake of saturated fatty acids, fiber, calcium, and potassium. Scoring above the 60th percentile of this score was defined as meeting the criteria for having a "healthy diet," consistent with other definitions of healthy diet in prior literature.^25^

BMI was calculated at the baseline visit for all participants, as individuals’ measured weight divided by their height, squared (kg/m^2^). Participants with BMI <18.5 or >70.0 were excluded from the analysis, as were those <20 years of age at the visit, consistent with quality control measures employed by other literature examining the genetic variation of BMI and obesity.^26^

### Genetic Data

Genotyping, quality control, and imputation methods employed in the HCHS/SOL cohort have been described elsewhere previously.^27,28^ Briefly, DNA was extracted from individual blood samples and genotyped on the SOL HCHS Custom 15041502 B3 array (i.e., Illumina Omni2.5M array + 150,000 custom informative SNPs).^27^ Standard quality control filters were enacted and then imputed using 1000 Genomes Project phase 3 reference populations. Principal components (PCs) of genetic ancestry, with 1000 Genomes reference populations, were constructed to account for confounding by population stratification.

The PGS_BMI_ employed in this analysis was derived from effect sizes published in a European-ancestry genome-wide association study (GWAS) of BMI in almost 700,000 participants enrolled in the UK Biobank and in the GIANT consortium, and applied to genome-wide data from HCHS/SOL.^29^ Using SNPs that passed a minor allele frequency cutoff of 5% in HCHS/SOL, the PGS_BMI_ was calculated from effect sizes in this GWAS via the LDpred method and an infinitesimal model.^30^ Several PGSs were compared to identify the best-fitting PGS_BMI_ which explained 7.4% of the variance in inverse normalized BMI in the HCHS/SOL cohort, adjusted for various relevant covariates. For consistency of interpretation, this PGS_BMI_ was then standardized to a mean of 0 and a standard deviation of 1.

### Genetic ancestry estimation

Genetic ancestry was estimated using genotyped data that passed the above quality control filters using a larger sample as detailed in a previous publication including other studies participating in the Population Architecture using Genomics and Epidemiology (PAGE) Study Genotype data were phased with SHAPEIT2 and imputed into 1000 Genomes phase 3 reference data using IMPUTE version 2.3.2.^31^ Unrelated individuals were subset from the data up to 2^nd^ degree relatives (N=45,255) and genotyped sites were pruned using r^2^<0.1 (M=132,591) in PLINK.^32^ Ancestry proportions were estimated using ADMIXTURE with 10 unsupervised runs assuming k=5.^13^ These five clusters were inferred to represent European (EUR), African (AFR), Amerindigenous (AME), East Asian (EAS), and Pacific Islander (PI) genetic ancestries given their distributions within self-identified racial and/or ethnic categories. Across all ten runs, there was only one mode as determined by pong (https://github.com/ramachandran-lab/pong), indicating stable estimates.^13,33^ Proportion of AME ancestry was expressed as a continuous value assigned to all participants within this HCHS/SOL sample (N=7,282).

### Statistical analysis

An exploratory analysis was conducted to characterize the variables that were most conceptually relevant to the model constructed herein, based on those chosen by McArdle *et al*. and described above. Table 1 p-values were calculated through the Wilcoxon rank-sum test for complex survey samples, which tests whether the group values are different from one another and via Pearson’s Chi-squared test with Rao & Scott’s second-order correction for categorical variables, which tests differences between observed and expected frequencies.

**Table 1.**
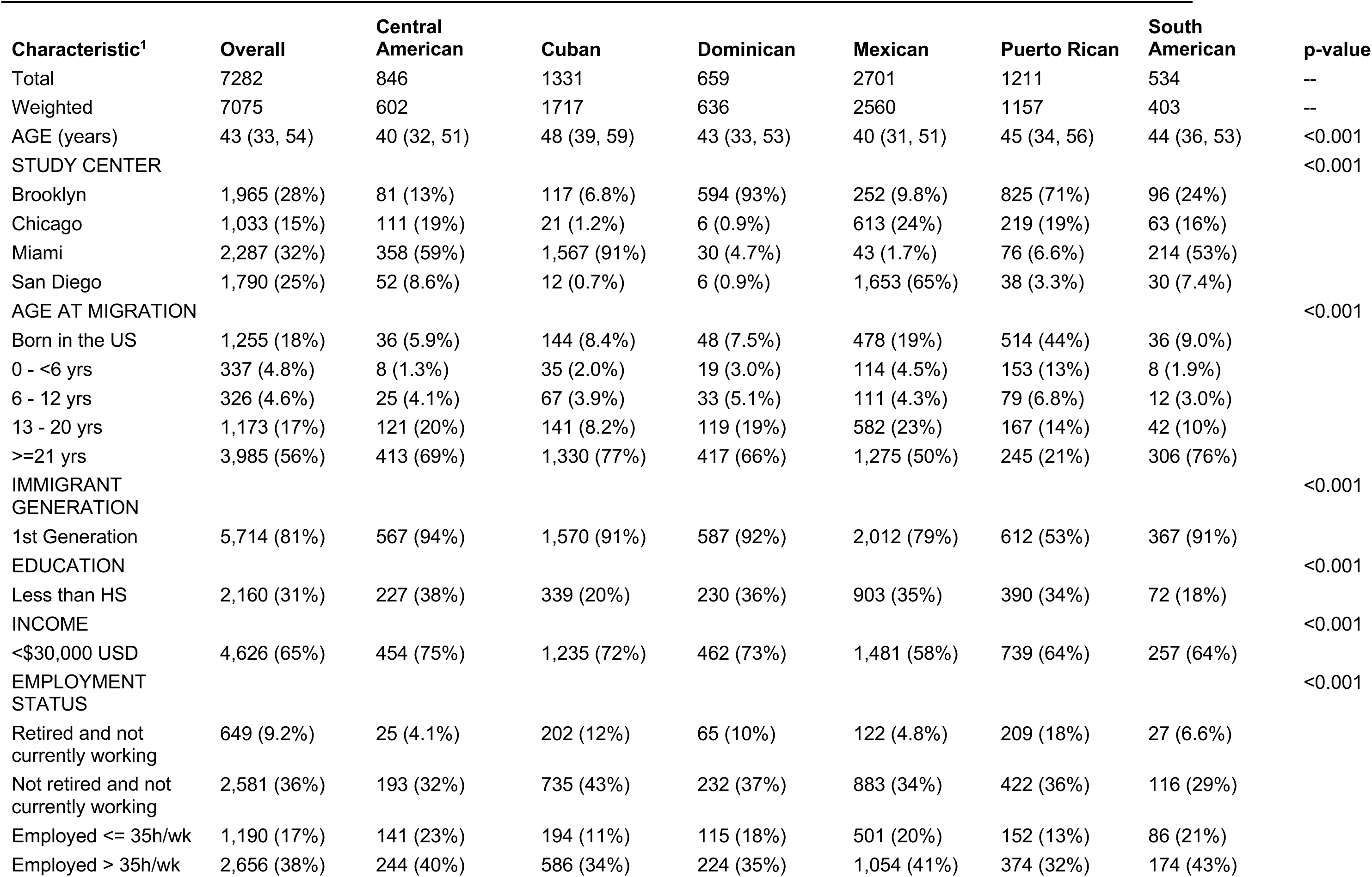
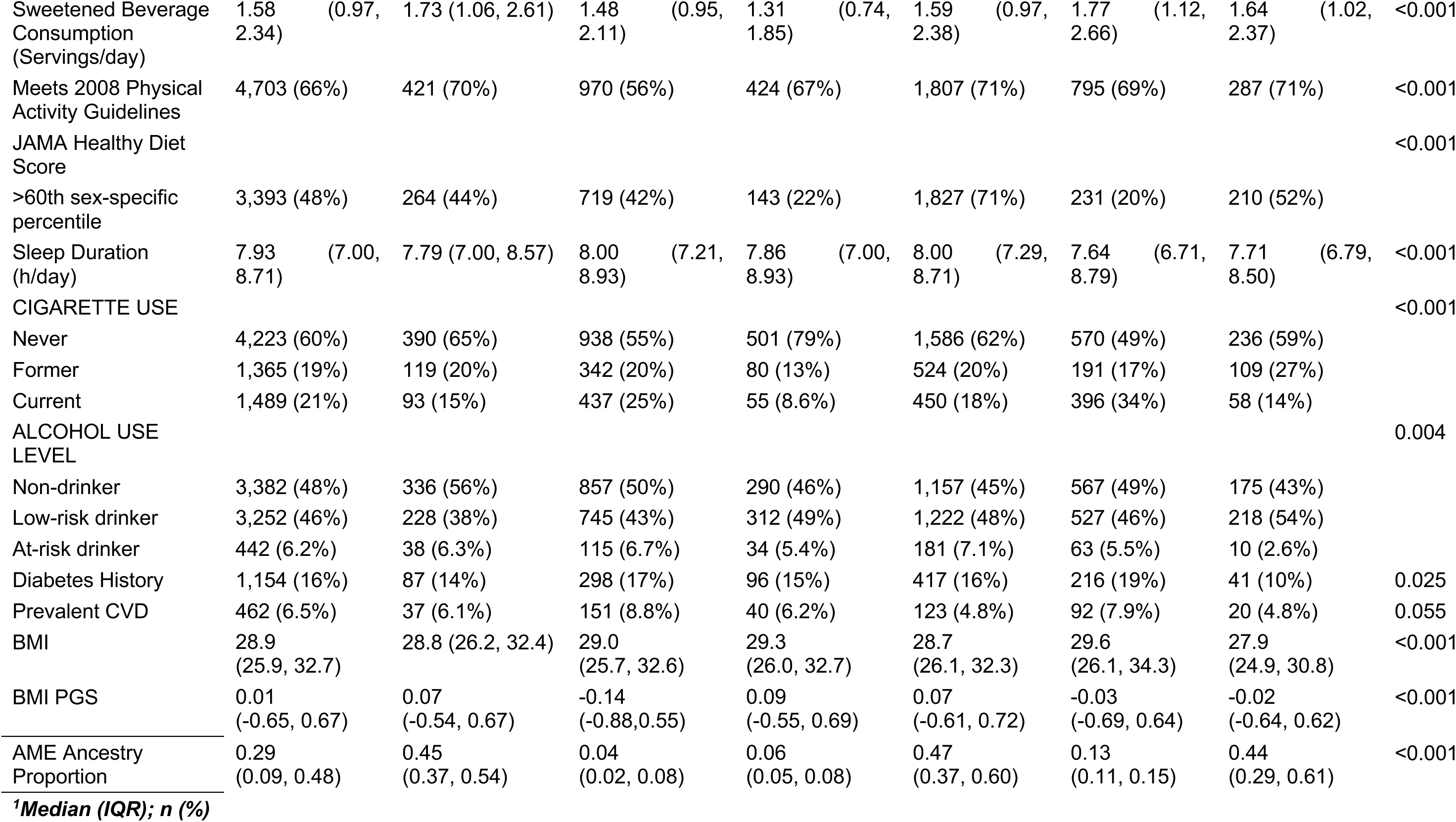
Selected Population Characteristics of the HCHS/SOL Analytic Subsample Overall (n=7,282) and Stratified by Background.

Models and observations used in summary measures were weighted using complex survey weights specified by HCHS/SOL protocols and all analyses were conducted in R version 4.1.1. Associations with proportion of AME ancestry were calculated through univariate survey-weighted generalized linear models of AME ancestry proportion on each individual explanatory variable included in Table 1, using the {gtsummary} package in R.^34^

To assess the contribution of AME ancestry proportion to the relationship between immigration-related variables, diet, and other measures as predictors of BMI, a generalized linear model was fit to the data comprising immigration-related and environmental variables as specified a priori by McArdle et al. The original model creation strategy used augmented backwards elimination and tests of GxE interaction that contributed to the development of a full model to predict BMI, and excluded variables such as SASH Language Scale, marital status, CESD-10 Depression Scale. The full model implemented in this analysis is shown below:

> *BMI ∼ PGS_BMI_ + 5 Principal Components (PCs) + Center + Age + Age^2^ + Sex + Diabetes + Sleep Duration + Cigarette Use + Physical Activity + CVD + Alcohol Use + Sweetened Beverage Consumption + Immigrant Generation + Employment Status + Education + Income + 5-category Age at Immigration + Healthy Diet Score + PGS_BMI_ *Healthy Diet Score + PGS_BMI_ *Age at Immigration*

To understand the influence of both ancestry and Hispanic/Latino background group, three modeling comparisons were employed: 1) implementing the full model with and without an AME ancestry proportion term, 2) stratifying the full model by quartile of AME ancestry, and 3) stratifying the full model by both group and by tertile of AME ancestry, since quartile stratification yielded too few observations in each stratification bin.

All models and observations used in summary measures were weighted using complex survey weights specified by HCHS/SOL protocols and all analyses were conducted in R version 4.1.1. Survey-weighted models were executed through the {survey} package in R.

## RESULTS

### Observed heterogeneity in key variables between Hispanic/Latino background groups

A total of 7,282 participants were included in the full analysis. After weighting to account for HCHS/SOL’s complex survey design and restricting to those with complete data for all included covariates, 7,075 weighted observations remained in the sample (representing 7,282 individuals). Detailed participant characteristics are described in Table 1. Within the weighted sample, individuals comprised six background groups: Central American (N=602), Cuban (N=1,717), Dominican (N=636), Mexican (N=2,560), Puerto Rican (N=1,157), and South American (N=403). We identified significant heterogeneity in population characteristics by background group, including BMI, immigrant generation, prevalent cardiovascular disease (CVD), and the PGS_BMI_. (Table 1) This difference is especially apparent between Caribbean and Central/South American geographical representations of Hispanic/Latino groups. Since some of the largest sample sizes are found in Mexican (N=2,560) and Puerto Rican (N=1,157) groups, and these two groups adequately demonstrate differences in geography by recruitment site and immigration-related histories, we will use these strata to illustrate these trends.

Overall, the average BMI across all Hispanic/Latino participants was 28.9 (IQR: 25.9, 32.7). However, the distributions of BMI were significantly different between Hispanic/Latino groups (p<0.001), with the mean BMI was slightly lower at 28.7 (IQR: 26.1, 32.3) in Mexican background individuals, while slightly higher in Puerto Rican background individuals at 29.6 (IQR: 26.1, 34.3). Significant heterogeneity was also observed when comparing genetic risk for BMI as estimated by the standardized PGS_BMI_ (p<0.001; Table 1). The weighted mean PGS_BMI_ in the pooled sample was 0.01 (IQR: -0.065, 0.067). However, the mean PGS_BMI_ was higher in the Mexican background group at 0.07 (IQR: -0.61, 0.72), and lower in the Puerto Rican background group at -0.03 (IQR: -0.69, 0.64), an inverse trend to BMI which showed lower values in Mexican background individuals and higher values in Puerto Rican background individuals.

Considering the main variables thought to interact with PGS_BMI_ and influence prediction of BMI (Figure 1), we note differences in proxy measures for acculturation used in this analysis. Overall, 80.8% of Hispanic/Latino individuals in this study identified as 1^st^ generation. However, when stratified, 79% of Mexican background participants identified as 1^st^ generation, compared to only 53% of Puerto Rican background individuals (Table 1). Additionally, there was significant heterogeneity in age-at-immigration between groups, with 50% of Mexican background participants immigrating after the age of 21 years old yet this proportion was only 21% of Puerto Rican background participants. Diet is another culture- and region-specific variable that demonstrates sociocultural differences. Only 29% of Mexican background individuals were below the 60th sex-specific percentile of their JAMA Healthy Diet score (less healthy diet), compared to 80% of Puerto Rican background individuals. This difference is not adequately depicted in the overall distribution of diet scores, where 52% of members of Hispanic/Latino ethnicity score below their sex-specific 60th percentile. Overall, this demonstrates marked heterogeneity in these key variables between Hispanic/Latino backgrounds which would be typically modeled as a single homogenous groups in genetic studies.

**Figure 1.**
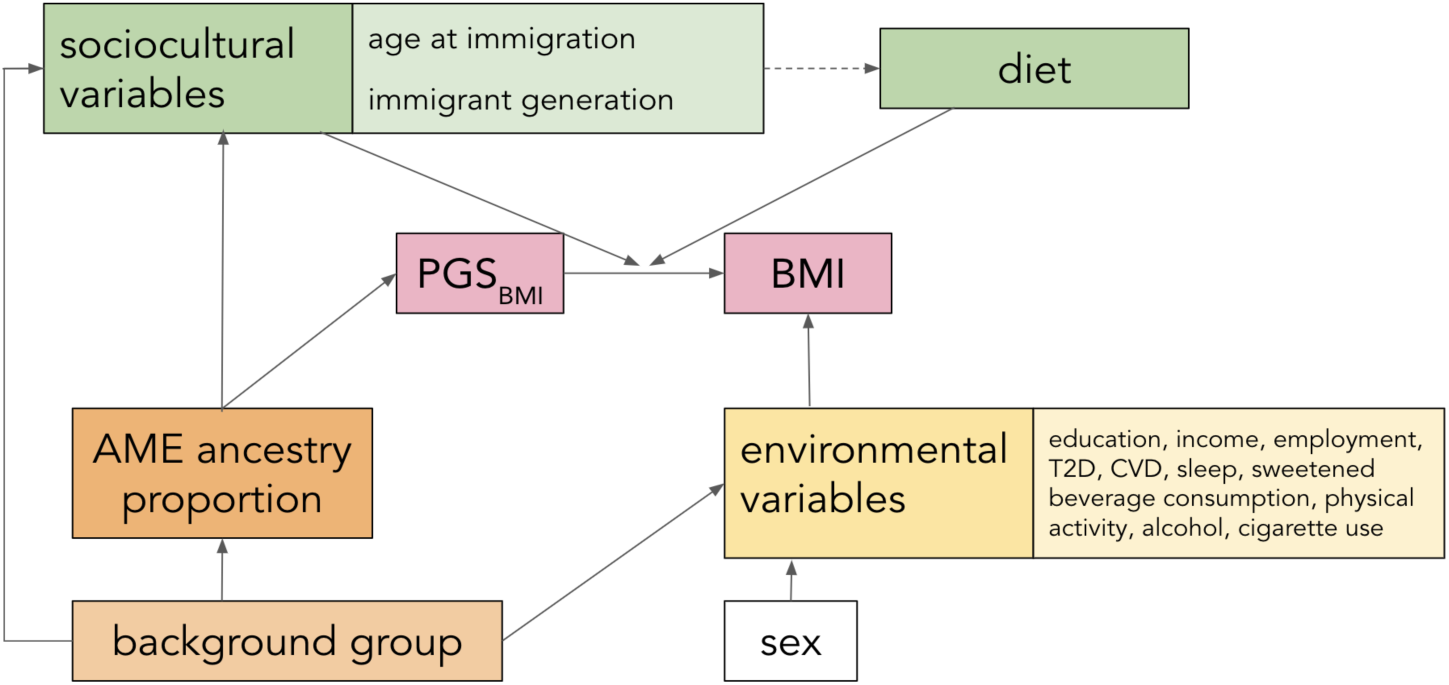
Conceptual Framework of Full Analytic Model.

### Relationships between inferred ancestry components with risk factors and traits differ by Hispanic/Latino background groups

In line with our conceptual framework (Figure 1), genetic ancestry, especially inferred AME ancestry in Hispanic/Latino ethnic groups, can capture finer genetic background composition in addition to self-identified background group that may confound the association of interest between PGS_BMI_ and BMI.^38^ We estimate differences in AME ancestry proportion distributions by Hispanic/Latino background group, and their associations with other risk factors (Table 2, Figure 2). While the pooled sample has on average 29% AME ancestry with noted variance (IQR: 9%, 48%), when stratified by background group reflecting geography and historical patterns of migration and colonialism, stark differences arise. Central American, Mexican, and South American background groups have higher mean AME ancestry of 45%, 44%, and 47%, respectively, while Cuban, Dominican, and Puerto Rican background groups, have lower mean AME ancestry proportions of 4%, 6%, and 13%, respectively (Supp Fig 1).

**Figure 2.**
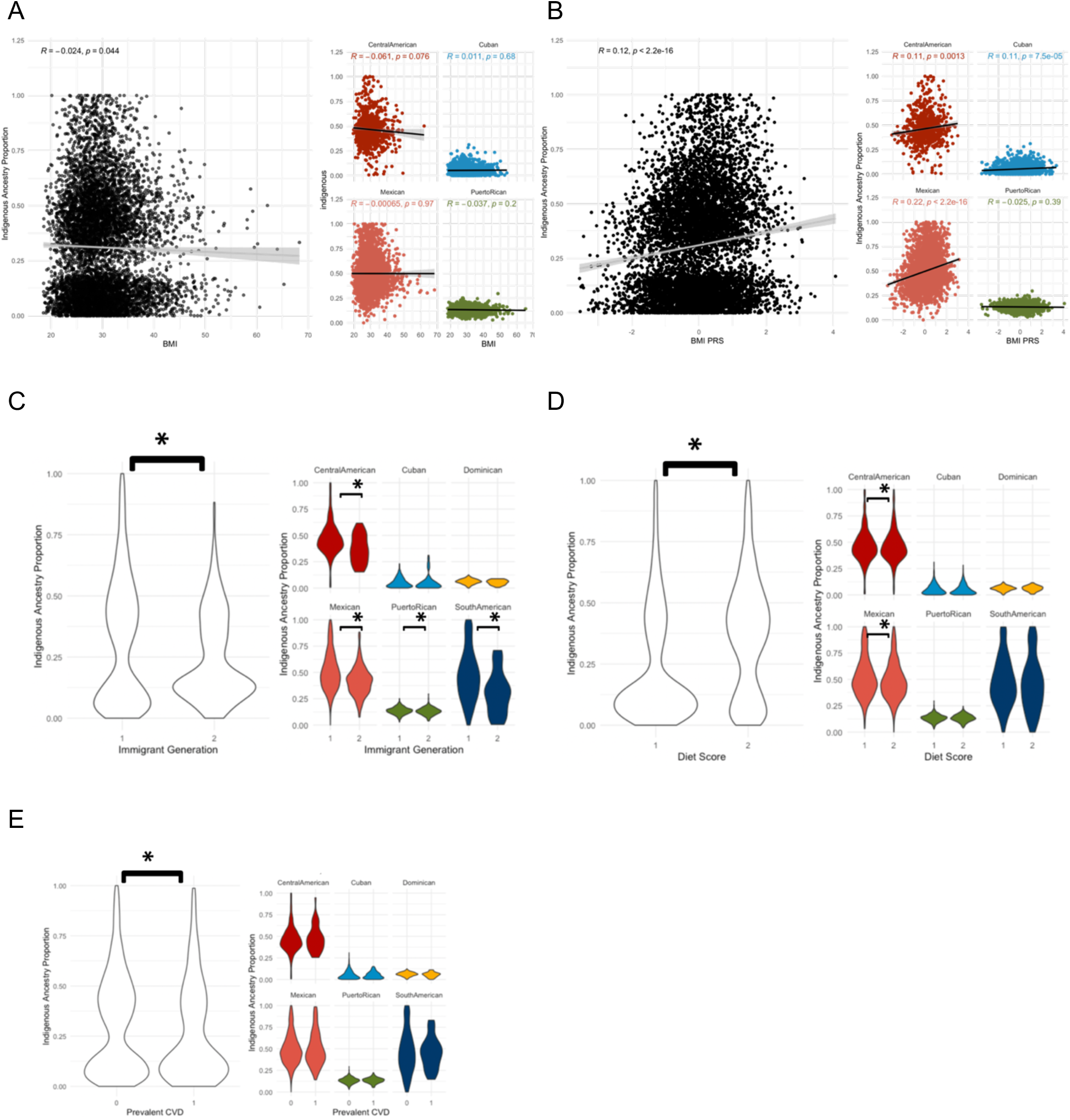
Distributions of Selected Variables by AME Ancestry Proportion Aggregated and by Background. Graphical depictions of relationships between AME ancestry proportion and A) BMI, B) PGS_BMI_, C) immigrant generation, D) diet score (1= under 60th sex-specific percentile of JAMA Healthy Diet score, 2= top 40th sex-specific percentile of JAMA Healthy Diet score), and E) prevalent cardiovascular disease in the complete sample (left panels; 1=yes, 0=no) and stratified by self-identified background group identity (right panels). * indicates statistically significant differences (p<0.05) between categories.

**Table 2.**
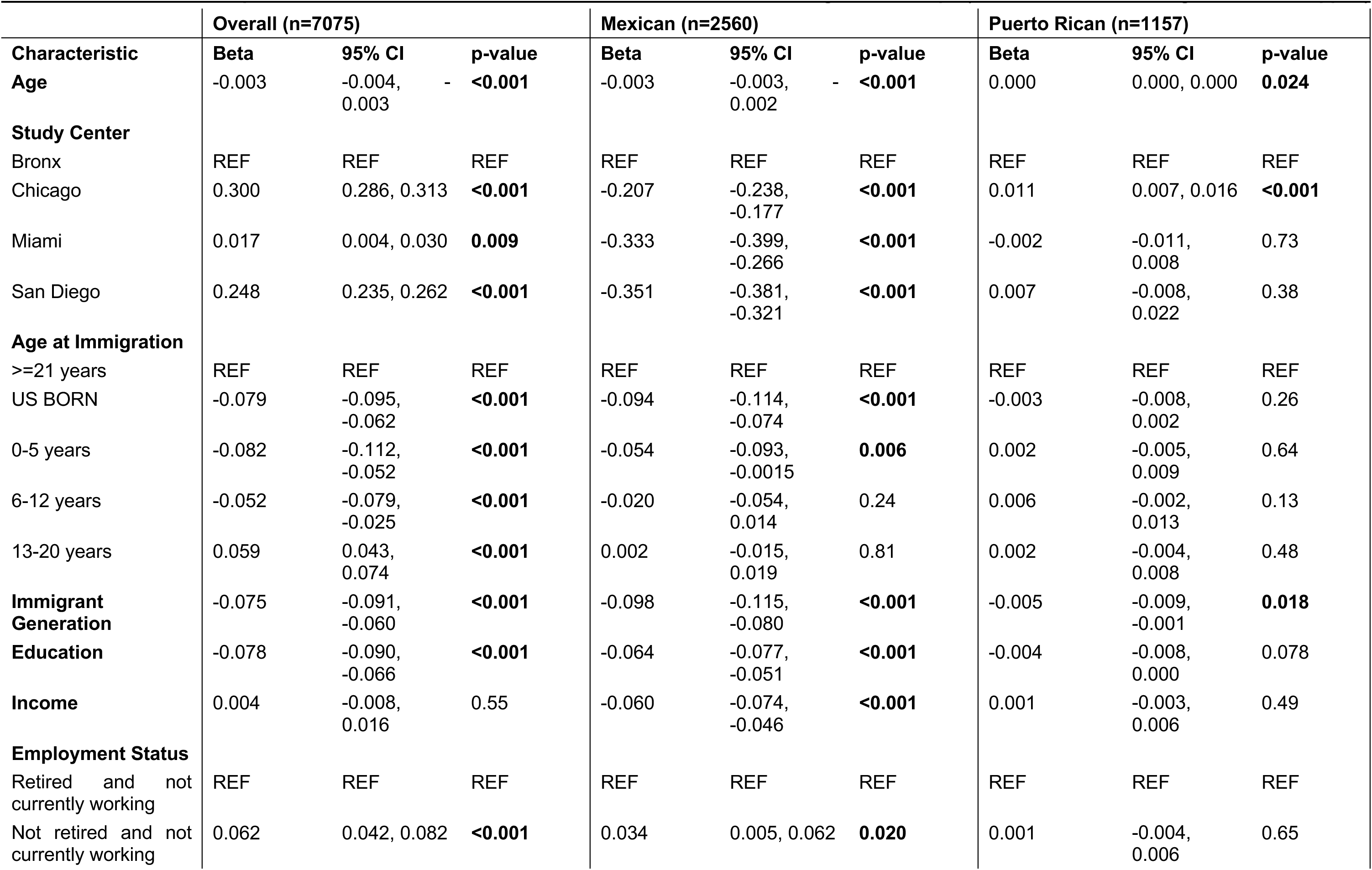
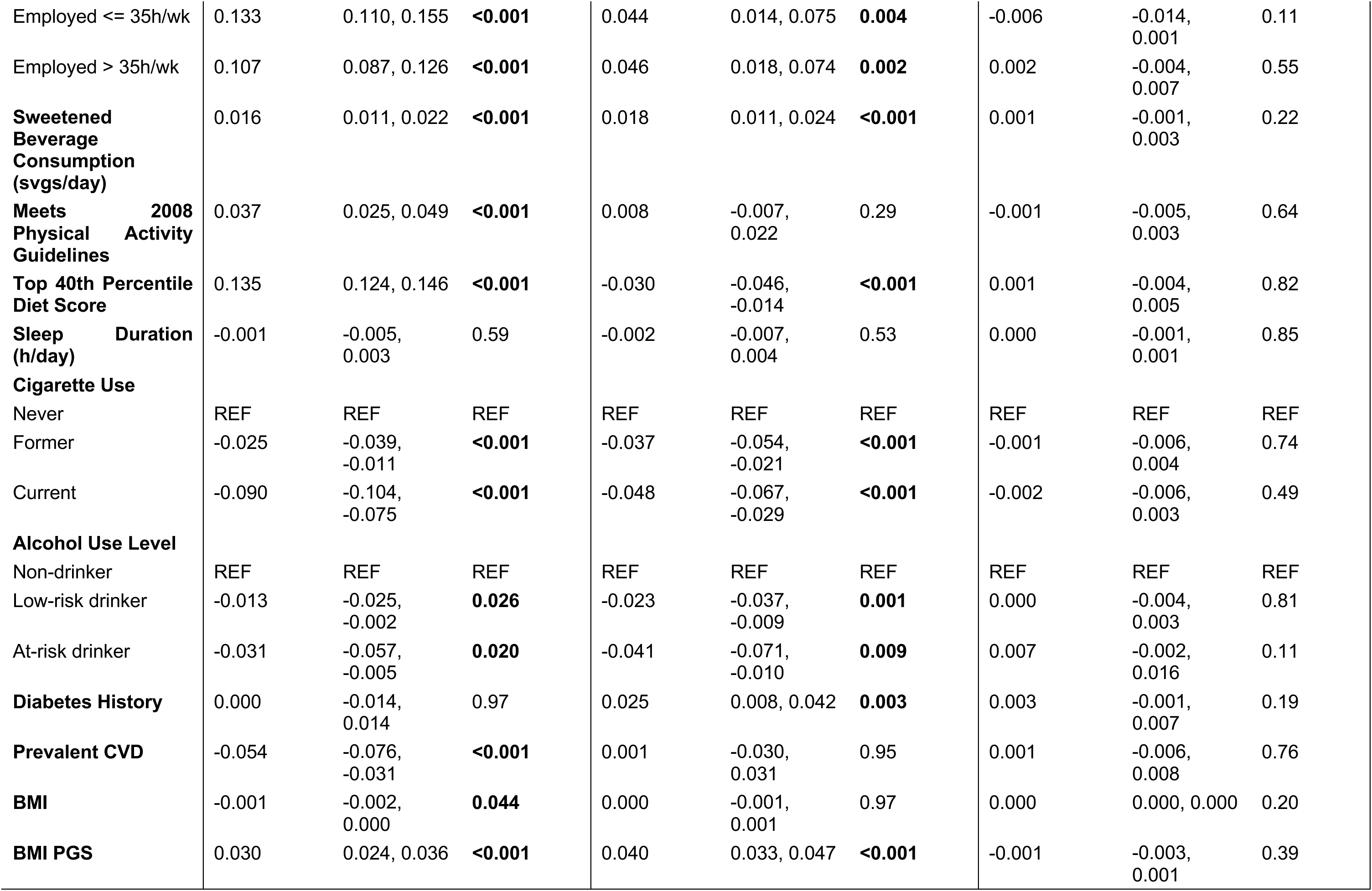
AME Ancestry Proportion Univariate Association with Selected Variables and Background Groups (Individuals with missing data were dropped)

To assess the relationship between these key variables and global ancestry proportions both in aggregate and between Hispanic/Latino groups, we conducted a univariate analysis between each variable and estimated proportion of AME ancestry. We observed similar heterogeneity to what was described above, largely driven by substantial AME ancestry distribution differences between background groups (Table 2). Specifically, we observed the association of AME ancestry to be significantly negatively associated with BMI among all participants (β=-0.001, p=0.044), but that effect is null when examined stratified by group (Figure 2A). When taken in conjunction with group-level differences in BMI as detailed above, this lack of statistically significant relationship after stratification indicates possible confounding of the relationship between ancestry and BMI by group membership.

Associations between the PGS_BMI_ and AME ancestry are similarly heterogeneous. Overall, there is a statistically significant positive association (β=0.030, p<0.001) among HCHS/SOL Hispanic/Latino participants. However, this association differs by background group with a larger effect size among Mexican background individuals (β=0.040, p<0.001), but attenuated signals in Puerto Rican background individuals (β=-0.001, p=0.39) (Figure 2B). In contrast to BMI, this indicates that the association between ancestry and PGS distribution is not spurious, but stronger in specific subgroups with higher AME ancestry proportions.

We observe significant heterogeneity in the association of AME ancestry proportions with other relevant variables, including prevalent cardiovascular disease (CVD) (Figure 2). The relationship between AME ancestry and CVD across the entire cohort appears statistically significant (β=-0.054 (95% CI: -0.076,-0.031), p<0.001). Importantly, when stratified by background group, this statistical significance disappears completely in all groups (p>0.05). This is consistent with our observation that certain groups have higher prevalence of CVD than others (e.g., Puerto Rican individuals at 7.9% vs. Mexican individuals at 4.8%), and that AME proportions vary widely by background group as well, indicating confounding by background group. Similar inferences are made regarding the Employment Status, Physical Activity, Marital Status, and CESD-10 Item Depression Score variables (Table 2). Taken together, these results caution the use of a pooled Hispanic/Latino participant sample to characterize the role of ancestry proportions on human health or relevant trait distributions.

### Interaction of PGS_BMI_ with non-genetic factors and admixture proportions in the pooled sample display heterogeneity

Given high heterogeneity in the univariate analyses, we sought to understand the influence of this heterogeneity in a model incorporating the above variables into a genetic risk prediction model for BMI. Specifically, to further investigate if differential performance of the PGS_BMI_ is reflective of potential population stratification not captured by principal components accounting for global genetic ancestry, we compared the performance of the extended model for BMI previously employed by McArdle *et al.* with and without an independent AME ancestry proportion term. In the pooled sample of all Hispanic/Latino participants, the addition of AME ancestry proportion to the full model did not meaningfully change the main effect of PGS_BMI_ on BMI (β=2.47, p<0.001 vs. β=2.46, p<0.001) or model fit with an R^2^ of 0.170 in the original full model from McArdle et al. and an R^2^ of 0.171 with the addition of AME ancestry proportions (Supp Table 1). Coefficients for other terms were consistent between models, including interaction terms with PGS_BMI_ with healthy diet and with age at immigration (Supp Table 1).

We then explored how proportion of AME ancestry may affect both the individual coefficients and overall model fit by stratifying the full model by inferred AME ancestry into quartiles in the pooled sample (Q1: 0-8.9%, Q2: 8.9-28.9%, Q3: 28.9-48.2%, and Q4: 48.2-99.9% AME ancestry). In contrast to the previous analysis which seeks to control for confounding, this analysis assesses the role of overall genetic background (AME ancestry proportion) on the ability for the PGS to capture the genetic liability for BMI. Overall, model fit (R^2^) generally decreased with increasing quartiles of AME ancestry (0.222, 0.202, 0.213, and 0.164, respectively; Table 3). Due to the collinearity of increasing inferred AME with decreasing EUR, this finding is consistent with prior literature showing generally decreased performance with increasing genetic distance from the training sample, which was of European ancestries.^40^ The adjusted effect size of the PGS_BMI_ on BMI remained generally consistent with increasing quartile of AME ancestry (quartile 1: β=2.23 (1.32,3.14), p<0.001; quartile 4: β=2.40 (1.27,3.52), p<0.001) (Supp Table 2).

**Table 3.** Comparing R^2^ values of multivariable regression results, stratified by AME ancestry proportion quartile.

| AME Ancestry Proportion |  | R<sup>2</sup> |
| --- | --- | --- |
| Quartile | Range |  |
| Quartile 1 | (0.000, 0.089) | 0.222 |
| Quartile 2 | (0.089, 0.289) | 0.202 |
| Quartile 3 | (0.289, 0.482) | 0.213 |
| Quartile 4 | (0.482, 0.999) | 0.164 |

Further, we examined the role of ancestry proportion on the effect modification of PGS_BMI_ by age-at-immigration and diet, which revealed heterogeneity by AME ancestry proportion quartile. The estimates of PGS_BMI_ x age-at-immigration interaction comparing those born in the US to immigrating as adults (>21 years) generally became stronger with increasing quartile of AME ancestry for the lowest three quartiles, though only statistically significant in the third quartile of AME ancestry (quartile 3: β=0.788 (95% CI: 0.183,1.29), p=0.01). The estimates of the PGS_BMI_-diet interaction were variable and only significant (p<0.05) in the highest quartile of AME ancestry (quartile 4: β=-0.536 (95% CI: -1.05, -0.03), p=0.04) (Supp Table 2). These interactions are directionally consistent with their unstratified complete model estimates for the PGS_BMI_-diet interaction (β=-0.398 (95% CI: -0.725, -0.07), p=0.017) and the PGS_BMI_-immigration interaction for adults >21 years compared to those born in the US (β=0.514 (95% CI: -0.126,1.15), p=0.12) (Supp Table 1).

### Interaction of PGS_BMI_ with non-genetic factors and admixture proportions differs by Hispanic/Latino background group

To evaluate the potential modification of both AME ancestry and background group on the performance of PGS_BMI_ and its interaction with sociocultural factors: we partitioned AME ancestry into tertiles in the pooled sample due to limited sample size when data is parsed by both variables. The ancestry tertiles corresponded to 0-12.5%, 12.5-42.8%, and 42.8-99% inferred AME ancestry. When stratified by both tertiles of AME ancestry and by background, model R^2^ decreased with increasing tertiles in all background groups with appreciable AME ancestry (Mexican, Central American, and South American). In Caribbean (Cuban, Dominican, Puerto Rican) groups with lower AME ancestry proportions on average, trends were inconsistent with insufficient data in some cells (Table 4). The adjusted effect of the PGS_BMI_ on BMI generally became stronger with increasing AME ancestry tertile in each background group, contrary to the trend observed without background stratification (Supp Tables 3.1-3.6). This trend was observed in all groups except in the Mexican background group, where the PGS_BMI_ coefficient became attenuated in the higher AME ancestry tertile (β=1.21 (95% CI: 0.190, 2.22), p=0.02) compared to the lower tertile (β=3.25 (95% CI: 1.72, 4.77), p<0.001).

**Table 4.** Comparing R^2^ values of multivariable regression results, stratified by AME ancestry proportion tertile & background group.

| AME Ancestry Proportion |  | Background Group (R<sup>2</sup> (n)) |  |  |  |  |  |
| --- | --- | --- | --- | --- | --- | --- | --- |
| Tertile | Range | Central American | Cuban | Dominican | Mexican | Puerto Rican | South American |
| 1 | (0.000, 0.125) | N/A (14) | 0.238 (1225) | 0.206 (658) | N/A (17) | 0.298 (476) | N/A (38) |
| 2 | (0.125, 0.428) | 0.295 (360) | 0.420 (106) | N/A (1) | 0.252 (1012) | 0.277 (735) | 0.351 (213) |
| 3 | (0.428, 0.999) | 0.220 (472) | N/A (0) | N/A (0) | 0.156 (1672) | N/A (0) | 0.333 (283) |

The estimates of the PGS_BMI_-immigration interaction (reference group are participants who immigrated at 21 years old or older) were only statistically significant in three strata: 1) in the 1st tertile of AME ancestry in the Cuban background group, comparing those aged 0-5 years at immigration (β=2.20 (0.43, 3.98), p=0.02), 2) in the 1st AME ancestry tertile of the Dominican background group, comparing those aged 6-12 years at immigration (β=4.38 (2.14, 6.62), p<0.001), and 3) in the 2nd AME ancestry tertile of the Mexican background group, comparing those born in the US (β=1.33 (0.59, 2.07), p<0.001). (Supplementary Figure 2) Expanding on the last strata for Mexican background individuals, participants who immigrated at ≥ 21 years old had on average an increase of 3.25 units of BMI (95% CI: 1.72, 4.77) for every standard deviation increase of PGS_BMI_. However, for participants born in the US, every one unit increase in PGS_BMI_ is associated with a 4.58 unit increase in BMI (1.33 units higher than the reference group). As such, the effect of the PRS is statistically higher in this contrast. The PGS_BMI_-immigration interaction remained statistically insignificant and was directionally heterogeneous in all other categories of background groups and subsequent ancestry tertiles, despite larger sample sizes in those bins. This indicates that even when explicitly controlling for ancestry, through both AME ancestry proportions and principal components, there remain differences between Hispanic/Latino groups for this GxE interaction (PGS_BMI_-immigration).

Beyond the inherent heterogeneity in model fit demonstrated here between background groups of a Hispanic/Latino cohort, we observe that AME ancestry proportion thereby differentially affects the GxE interactions underlying BMI-diet-age at immigration relationships. As AME ancestry proportion increases, the model fit worsens. Interestingly, the statistically significant GxE interactions of PGS-by-diet and PGS-by-age at immigration generally did persist upon stratification by ancestry quartile and by both ancestry tertile and background, except in the 1st tertiles of Mexican and Dominican groups and in the 2nd tertile of the Central American background group, where estimates were directionally heterogeneous between groups. This effect size heterogeneity across ancestry tertiles within some groups may indicate the intersectional role of genetic ancestry with sociocultural influences which could affect the performance of the PGS.

## DISCUSSION

Through the present analyses, we sought to characterize these complex relationships between a polygenic score, genetic ancestry, background group, and various lifestyle, sociodemographic, and immigration-related variables in BMI prediction in a Hispanic/Latino population (HCHS/SOL). Our inquiry was four-fold: (1) what are the individual distributions of these variables across Hispanic/Latino backgrounds?; (2) what is these variables’ relationship to genetic ancestry?; (3) can inferred genetic ancestry explain previously identified heterogeneity between Hispanic/Latino backgrounds in an integrated BMI risk model?; and (4) does additional self-identified background group-level stratification offer additional insights to comprehensive inferred ancestry modeling? Across all analyses, we identified persistent heterogeneity in univariate and multivariate associations of these variables with PGS_BMI_, particularly due to confounding by Hispanic/Latino background group that indicates the importance of background-level stratification in avoiding spurious findings in this population.

First, we identified high levels of heterogeneity among the various genetic and environmental variables in this study population, underscoring the important distinctions to be made between groups of self-identified Hispanic/Latino ethnicity. In fact, aggregating groups of this ethnicity obscures meaningful heterogeneity between geographical and cultural groups of Hispanic/Latino ethnicity and may lead to false conclusions. This is consistent with other studies of acculturation and BMI in Hispanic/Latino populations. For example, Khan *et al*. found that the effect of acculturation on BMI among Mexican Americans in the Hispanic Health and Nutrition Examination Survey was stronger than among Puerto Ricans and Cubans, which the authors hypothesized as being at least partly a function of longer duration of "exposure to (mainland) US culture" among Mexican Americans, and may have also been driven by a higher sample size in this group.^10^

Second, we observed that when stratified by background group, the relationships of these variables with genetic ancestry, modeled here as inferred AME ancestry admixture proportion, were heterogeneous. Genetic analyses of Hispanic/Latino participants may be inclined to incorporate Amerindigenous or other ancestry proportions into the model, as an important predictor of environmental variables of interest. It has been reported that those of Hispanic/Latino ethnicity in the US have, on average, 18.0% AME ancestry.^35^ The analysis of AME ancestry in this cohort, however, demonstrates significant variation ranging from an average of 4% to 47% AME ancestry in different Hispanic/Latino background groups. In our analysis, we identify heterogeneity in associations between various health-related variables and ancestry proportions, both between pooled and stratified results, as well as between the groups themselves. Therefore, in studies involving Hispanic/Latino participants, background group stratification is important to prevent inferences of spurious associations of such variables with AME ancestry, which are instead driven primarily by group heterogeneity and differences in ancestry proportions between background groups.

Third, we found that the fit of our predictive model for BMI as explained by a PGS_BMI_ along with various health, immigration, and environmental variables, and genetic ancestry worsened with increasing proportions of AME ancestry. This observation is consistent with previous demonstrations of poor transferability of PGS trained in European-ancestries to other populations.^36,37^ This is not surprising given of the PGS_BMI_ here was trained on a largely European ancestry sample, which inherently does not reflect the genetic composition of HCHS/SOL participants.^29^ However, it is notable that there is significant heterogeneity within the pooled Hispanic/Latino participants. Longstanding patterns of migration, conflict, and other sociopolitical factors influenced the current structure of genetic ancestry of Hispanic/Latino groups being a combination of European, African, and Amerindigenous ancestry, giving rise to different allele frequencies and linkage disequilibrium (LD) correlation structures.^35^ This poor performance by ancestry has consequences in the ability to detect GxE interactions, as demonstrated by the high heterogeneity in interaction effect estimates of PGS with immigration and diet-related variables, particularly related to the confounding effect of background group on the relationship of these variables and BMI.

Fourth, when we subsequently stratified this model by both AME ancestry and background group, we identified heterogeneous effects in the specific contrasts in which GxE interactions persisted. McArdle *et al*. previously identified few statistically significant GxE interactions in an analysis of the full model stratified by background group: those who immigrated to the US had a higher BMI than those born in the US, comparing those with a higher to lower PGS_BMI_.^21^ In our analysis that was stratified by *both* background group and AME ancestry, our results also indicated a modest effect of younger age at immigration to the US on the PGS_BMI_- BMI relationship in Cuban, Dominican, and Mexican background groups. Those who were brought to the US at young ages from different countries may also reflect different demographic backgrounds and sociocultural environments that represent varying risk-increasing or risk-decreasing relationships of health- or environment-related risk factors with BMI and obesity. Group-specific histories of colonization, along with geopolitical and temporal patterning of how specific Hispanic/Latino groups became part of or relocated to the US, play a major role in these sociocultural and demographic differences, with immigration and land loss beginning in the mid-19^th^ century and continuing to the present day through many immigration barriers and policies.^38^ In recent history, migration from Mexico to the US peaked between 2000-2010, before the implementation of stricter border policies that curtailed the routine back and forth or "circular" migration patterns between Mexico and the US.^39,40^ The Caribbean islands of Puerto Rico, the Dominican Republic, and Cuba, despite their geographical proximity, have had more nuanced US migration and immigration patterns. While the former was annexed by the US in 1898 and saw migration numbers peak after World War II, Cubans were mostly isolated from the US during this time.^39^ This long standing relationship changed, however, with the 1959 communist revolution, which drove the immigration of hundreds of thousands of Cuban migrants to the US.^39^ In recent years, the immigration of Hispanic/Latino people to the US has continued, including many from Central and South American countries, albeit at a slower pace in the face of stricter immigration policy.^41^ Our results are at least partially consistent with other non-genetic analyses of obesity in Hispanic/Latino populations in the US that reflect the important role of US residency and immigration in obesity and that age at immigration is not as significant an obesity risk factor in these populations.^42^

This study’s limitations lie primarily in small sample size and potential for information bias and misclassification. Loss of statistical significance in the GxE interactions is likely driven at least in part by small sample sizes in the increasingly minute sub-classifications of the analytic sample. However, the maintained statistical significance of these interactions and their significant effect size change in two of the background groups suggests that group analyses could identify differential modifying effects of diet and age-at-immigration on the effect on BMI of PGS_BMI_. Our analyses were also restricted to incorporation of AME ancestry, despite the notably important contribution of African (AFR) ancestry, particularly in Dominican background participants. High AFR ancestry likely also contributes differentially to poor model fit observed for this group in Table 5.^43^ We did not explore associations of immigration-related, environmental, and diet variables with African (AFR) ancestry, due to issues arising from collinearity of AFR ancestry with AME ancestry and extremely limited sample sizes poorly powered to detect effects. Further studies to examine the role of AFR ancestry in these GxE frameworks are needed.

An additional limitation is that the measurement of immigration-related variables (i.e. age-at-immigration, nativity, and years lived in the US) requires the simplification of innately complex concepts that reflect years and often generations of migratory and environmental changes. Importantly, these analyses cannot capture the circular nature of migration of Hispanic/Latino groups, for example among individuals of Mexican and Puerto Rican backgrounds who often migrate between the US 50 states/DC and Mexico or Puerto Rico within and across generations.^39,40,44,45^ However, current analyses of health and health-related behaviors in US Hispanic/Latino populations have consistently used acculturation and immigration-related variables as proxies for complex migratory and environmental patterns and have robustly identified associations with health-related variables, including BMI.^46–48^

Despite controlling for self-identified background, AME ancestry proportion, and principal components, our analyses still identified significant heterogeneity in GxE interactions, indicating the influence of non-genetic factors and complex social environments in the poor performance of a model exploring a European-ancestry-derived PGS_BMI_ in a non-European sample. Taken together, we demonstrate that environmental variables play an important role in the effect of genetics on obesity risk in Hispanic/Latino groups. However, the consideration of group identity within Hispanic/Latino ethnicity and other large ethnic groups requires increased urgency, especially as the incorporation of precision medicine into clinical practice and preventative care looms closer. Heeding calls for movement towards an integrated risk score requires acknowledgement of the complex genetic and environmental profiles of this population, specifically the explicit modeling of their multi-layered sociodemographic and relationally complex migration histories of which may confound relationships of genetic and environmental variables.

To this aim, we recommend that cohorts must collect finer data on Hispanic/Latino participants that will continue to be used to study this population. This recommendation falls in line with recent consensus around for the appropriate use of race and ethnicity as population descriptors in both genetic and biomedical research: to use more granular levels of participants’ self-identity rather than ancestry-driven or broadly defined categorization.^49,50^ Additionally, clinical applications of genetic association analyses and of analyses incorporating gene-environment interactions should pay attention to different modifying effects of environmental variables between Hispanic/Latino background groups. More broadly, epidemiologic and other analyses should stratify Hispanic/Latino participant populations by their background group identity to understand the true nature of public health relationships in these groups, and to prevent the publication of sweeping generalizations that are not reflective of true associations between important variables.

## Data Availability

All data produced in the present study are available upon reasonable request to the authors.

## Acknowledgements

The authors thank the staff and participants of HCHS/SOL for their important contributions. (Investigators website - http://www.cscc.unc.edu/hchs/) The Hispanic Community Health Study/Study of Latinos is a collaborative study supported by contracts from the National Heart, Lung, and Blood Institute (NHLBI) to the University of North Carolina (HHSN268201300001I / N01-HC-65233), University of Miami (HHSN268201300004I / N01-HC-65234), Albert Einstein College of Medicine Rev. December 4, 2023 (HHSN268201300002I / N01-HC-65235), University of Illinois at Chicago (HHSN268201300003I / N01-HC-65236 Northwestern Univ), and San Diego State University (HHSN268201300005I / N01-HC-65237). The following Institutes/Centers/Offices have contributed to the HCHS/SOL through a transfer of funds to the NHLBI: National Institute on Minority Health and Health Disparities, National Institute on Deafness and Other Communication Disorders, National Institute of Dental and Craniofacial Research, National Institute of Diabetes and Digestive and Kidney Diseases, National Institute of Neurological Disorders and Stroke, NIH Institution-Office of Dietary Supplements. Additional funding for JS and GLW comes from the National Human Genome Research Institute (R35HG011944).

## REFERENCES

1. Ogden CL, Carroll MD, Fryar CD, Flegal KM. Prevalence of Obesity Among Adults and Youth: United States, 2011-2014. NCHS Data Brief. 2015;(219):1-8. https://www.ncbi.nlm.nih.gov/pubmed/26633046

2. Krogstad JM. Hispanics only group to see its poverty rate decline and incomes rise. Published September 19, 2014. Accessed August 7, 2024. https://www.pewresearch.org/short-reads/2014/09/19/hispanics-only-group-to-see-its-poverty-rate-decline-and-incomes-rise/

3. Cortes-Bergoderi M, Goel K, Murad MH, et al. Cardiovascular mortality in Hispanics compared to non-Hispanic whites: a systematic review and meta-analysis of the Hispanic paradox. Eur J Intern Med. 2013;24(8):791–799. doi:10.1016/j.ejim.2013.09.003

4. Chartier KG, Carmody T, Akhtar M, Stebbins MB, Walters ST, Warden D. Hispanic Subgroups, Acculturation, and Substance Abuse Treatment Outcomes. J Subst Abuse Treat. 2015;59:74–82. doi:10.1016/j.jsat.2015.07.008

5. Smith TM, Colón-Ramos U, Pinard CA, Yaroch AL. Household food insecurity as a determinant of overweight and obesity among low-income Hispanic subgroups: Data from the 2011-2012 California Health Interview Survey. Appetite. 2016;97:37–42. doi:10.1016/j.appet.2015.11.009

6. Manichaikul A, Palmas W, Rodriguez CJ, et al. Population structure of Hispanics in the United States: the multi-ethnic study of atherosclerosis. PLoS Genet. 2012;8(4):e1002640. doi:10.1371/journal.pgen.1002640

7. Gravel S, Zakharia F, Moreno-Estrada A, et al. Reconstructing Native American migrations from whole-genome and whole-exome data. PLoS Genet. 2013;9(12):e1004023. doi:10.1371/journal.pgen.1004023

8. US Census Bureau. Measuring Racial and Ethnic Diversity for the 2020 Census. Accessed March 17, 2022. https://www.census.gov/newsroom/blogs/random-samplings/2021/08/measuring-racial-ethnic-diversity-2020-census.html

9. Hales CM, Carroll MD, Fryar CD, Ogden CL. Prevalence of obesity and severe obesity among adults: United States, 2017-2018. NCHS Data Brief. 2020;(360):1-8. Accessed February 27, 2022. https://www.cdc.gov/nchs/data/databriefs/db360-h.pdf

10. Khan LK, Sobal J, Martorell R. Acculturation, socioeconomic status, and obesity in Mexican Americans, Cuban Americans, and Puerto Ricans. Int J Obes Relat Metab Disord. 1997;21(2):91–96. doi:10.1038/sj.ijo.0800367

11. Yracheta JM, Alfonso J, Lanaspa MA, et al. Hispanic Americans living in the United States and their risk for obesity, diabetes and kidney disease: Genetic and environmental considerations. Postgrad Med. 2015;127(5):503–510. doi:10.1080/00325481.2015.1021234

12. Tang H, Peng J, Wang P, Risch NJ. Estimation of individual admixture: analytical and study design considerations. Genet Epidemiol. 2005;28(4):289–301. doi:10.1002/gepi.20064

13. Alexander DH, Novembre J, Lange K. Fast model-based estimation of ancestry in unrelated individuals. Genome Res. 2009;19(9):1655–1664. doi:10.1101/GR.094052.109

14. Homburger JR, Moreno-Estrada A, Gignoux CR, et al. Genomic Insights into the Ancestry and Demographic History of South America. PLoS Genet. 2015;11(12):e1005602. doi:10.1371/journal.pgen.1005602

15. Moreno-Estrada A, Gravel S, Zakharia F, et al. Reconstructing the population genetic history of the Caribbean. PLoS Genet. 2013;9(11):e1003925. doi:10.1371/journal.pgen.1003925

16. Bryc K, Velez C, Karafet T, et al. Colloquium paper: genome-wide patterns of population structure and admixture among Hispanic/Latino populations. Proc Natl Acad Sci U S A. 2010;107 Suppl 2(Supplement_2):8954-8961. doi:10.1073/pnas.0914618107

17. Martin AR, Kanai M, Kamatani Y, Okada Y, Neale BM, Daly MJ. Clinical use of current polygenic risk scores may exacerbate health disparities. Nat Genet. 2019;51(4):584–591. doi:10.1038/s41588-019-0379-x

18. Bitarello BD, Mathieson I. Polygenic scores for height in admixed populations. G3 (Bethesda). 2020;10(11):4027-4036. doi:10.1534/g3.120.401658

19. Mostafavi H, Harpak A, Agarwal I, Conley D, Pritchard JK, Przeworski M. Variable prediction accuracy of polygenic scores within an ancestry group. Elife. 2020;9. doi:10.7554/eLife.48376

20. Tyrrell J, Zheng J, Beaumont R, et al. Genetic predictors of participation in optional components of UK Biobank. Nat Commun. 2021;12(1):1–13. doi:10.1038/s41467-021-21073-y

21. McArdle CE, Bokhari H, Rodell CC, et al. Findings from the Hispanic Community Health Study/Study of Latinos on the Importance of Sociocultural Environmental Interactors: Polygenic Risk Score-by-Immigration and Dietary Interactions. Front Genet. 2021;12:1853. doi:10.3389/FGENE.2021.720750/BIBTEX

22. LaVange LM, Kalsbeek WD, Sorlie PD, et al. Sample Design and Cohort Selection in the Hispanic Community Health Study/Study of Latinos. Ann Epidemiol. 2010;20(8):642–649. doi:10.1016/J.ANNEPIDEM.2010.05.006

23. World Health Organization 2020 Guidelines on Physical Activity and Sedentary Behaviour.

24. Andresen EM, Malmgren JA, Carter WB, Patrick DL. Screening for depression in well older adults: evaluation of a short form of the CES-D (Center for Epidemiologic Studies Depression Scale). Am J Prev Med. 1994;10(2):77–84. https://www.ncbi.nlm.nih.gov/pubmed/8037935

25. Liu K, Daviglus ML, Loria CM, et al. Healthy lifestyle through young adulthood and the presence of low cardiovascular disease risk profile in middle age: the Coronary Artery Risk Development in (Young) Adults (CARDIA) study. Circulation. 2012;125(8):996–1004. doi:10.1161/CIRCULATIONAHA.111.060681

26. Gong J, Nishimura KK, Fernandez-Rhodes L, et al. Trans-ethnic analysis of metabochip data identifies two new loci associated with BMI. Int J Obes. 2017;42(3):384–390. doi:10.1038/ijo.2017.304

27. Conomos MP, Laurie CA, Stilp AM, et al. Genetic Diversity and Association Studies in US Hispanic/Latino Populations: Applications in the Hispanic Community Health Study/Study of Latinos. Am J Hum Genet. 2016;98(1):165–184. doi:10.1016/j.ajhg.2015.12.001

28. Sofer T, Moon JY, Isasi CR, et al. Relationship of genetic determinants of height with cardiometabolic and pulmonary traits in the Hispanic Community Health Study/Study of Latinos. Int J Epidemiol. 2018;47(6):2059–2069. doi:10.1093/ije/dyy177

29. Yengo L, Sidorenko J, Kemper KE, et al. Meta-analysis of genome-wide association studies for height and body mass index in ∼700000 individuals of European ancestry. Hum Mol Genet. 2018;27(20):3641–3649. doi:10.1093/hmg/ddy271

30. Vilhjálmsson BJ, Yang J, Finucane HK, et al. Modeling Linkage Disequilibrium Increases Accuracy of Polygenic Risk Scores. Am J Hum Genet. 2015;97(4):576–592. doi:10.1016/j.ajhg.2015.09.001

31. Wojcik GL, Graff M, Nishimura KK, et al. Genetic analyses of diverse populations improves discovery for complex traits. Nature. 2019;570(7762):514-518. doi:10.1038/s41586-019-1310-4

32. Chang CC, Chow CC, Tellier LC, Vattikuti S, Purcell SM, Lee JJ. Second-generation PLINK: rising to the challenge of larger and richer datasets. Gigascience. 2015;4(1):7. doi:10.1186/s13742-015-0047-8

33. Behr AA, Liu KZ, Liu-Fang G, Nakka P, Ramachandran S. pong: fast analysis and visualization of latent clusters in population genetic data. Bioinformatics. 2016;32(18):2817–2823. doi:10.1093/BIOINFORMATICS/BTW327

34. Sjoberg DD. Gtsummary: Presentation-Ready Data Summary and Analytic Result Tables.; 2021. https://CRAN.R-project.org/package=MetBrewer

35. Bryc K, Durand EY, Macpherson JM, Reich D, Mountain JL. The genetic ancestry of African Americans, Latinos, and European Americans across the United States. Am J Hum Genet. 2015;96(1):37–53. doi:10.1016/j.ajhg.2014.11.010

36. Duncan L, Shen H, Gelaye B, et al. Analysis of polygenic risk score usage and performance in diverse human populations. Nat Commun. 2019;10(1):1–9. doi:10.1038/s41467-019-11112-0

37. Grinde KE, Qi Q, Thornton TA, et al. Generalizing polygenic risk scores from Europeans to Hispanics/Latinos. Genet Epidemiol. 2019;43(1):50–62. doi:10.1002/gepi.22166

38. Alba FAF. Mexico: The New Migration Narrative. Accessed March 31, 2022. https://www.migrationpolicy.org/article/mexico-new-migration-narrative

38. Blizzard JBB, Batalova J. Cuban Immigrants in the United States. Accessed February 27, 2022. https://www.migrationpolicy.org/article/cuban-immigrants-united-states-2018

40. Rosenblum MR, Kandel WA, Seelke CR, Wasem RE. Mexican migration to the United States: Policy and trends. Published 2012. Accessed March 18, 2022. https://sgp.fas.org/crs/row/R42560.pdf

41. Moslimani M. Facts on Latinos in the U.S. Published August 16, 2023. Accessed August 6, 2024. https://www.pewresearch.org/race-and-ethnicity/fact-sheet/latinos-in-the-us-fact-sheet/

42. Isasi CR, Ayala GX, Sotres-Alvarez D, et al. Is acculturation related to obesity in Hispanic/Latino adults? Results from the Hispanic Community Health Study/Study of Latinos. J Obes. 2015;2015. doi:10.1155/2015/186276

43. Tajima A, Hamaguchi K, Terao H, et al. Genetic background of people in the Dominican Republic with or without obese type 2 diabetes revealed by mitochondrial DNA polymorphism. J Hum Genet. 2004;49(9):495–499. doi:10.1007/s10038-004-0179-7

44. Duany J. Mobile livelihoods: The sociocultural practices of circular migrants between Puerto Rico and the United States. Int Migr Rev. 2002;36(2):355–388. doi:10.1111/j.1747-7379.2002.tb00085.x

45. Durand J, Massey DS. Evolution of the Mexico-U.s. migration system: Insights from the Mexican Migration Project. Ann Am Acad Pol Soc Sci. 2019;684(1):21–42. doi:10.1177/0002716219857667

46. Lara M, Gamboa C, Kahramanian MI, Morales LS, Bautista DEH. Acculturation and Latino health in the United States: a review of the literature and its sociopolitical context. Annu Rev Public Health. 2005;26:367–397. doi:10.1146/annurev.publhealth.26.021304.144615

47. Fernández-Rhodes L, Butera NM, Lodge EK, et al. Demographic and sociocultural risk factors for adulthood weight gain in Hispanic/Latinos: results from the Hispanic Community Health Study / Study of Latinos (HCHS/SOL). BMC Public Health. 2021;21(1):2064. doi:10.1186/s12889-021-11848-9

48. Guadamuz JS, Durazo-Arvizu RA, Daviglus ML, et al. Immigration Status and Disparities in the Treatment of Cardiovascular Disease Risk Factors in the Hispanic Community Health Study/Study of Latinos (Visit 2, 2014–2017). Am J Public Health. 2020;110(9):1397-1404. doi:10.2105/AJPH.2020.305745

49. National Academies of Sciences, Engineering, and Medicine. 2023. Using Population Descriptors in Genetics and Genomics Research: A New Framework for an Evolving Field. Washington, DC: The National Academies Press. 10.17226/26902.

50. National Academies of Sciences, Engineering, and Medicine. 2024. Rethinking Race and Ethnicity in Biomedical Research. Washington, DC: The National Academies Press. 10.17226/27913.

