## Supplementary Information for "Influence of Genetic Ancestry on Gene-Environment Interactions of Polygenic Risk and Sociocultural Factors: Results from the Hispanic Community Health Study/Study of Latinos"

#### SUPPLEMENTAL INFORMATION

Additional Information for Figure 1.
*Variables and relationships modeled in this conceptual framework reflect the modeling approaches undertaken in the present analysis. AME= inferred Amerindigenous ancestry; BMI= body mass index; CVD=cardiovascular disease; PGS=polygenic score; T2D=type 2 diabetes.*

Supplemental Table 1. Regression Coefficients comparing Models with and without AME Ancestry Proportion

| | **Model 1 (without AME Ancestry)** | | | **Model 2 (with AME Ancestry)** | | |
|---|---|---|---|---|---|---|
| **Characteristic** | **Beta** | **95% CI** | **p-value** | **Beta** | **95% CI** | **p-value** |
| $PGS_{BMI}$ | 2.47 | 1.59, 3.35 | <0.001 | 2.46 | 1.59, 3.33 | <0.001 |
| PC1 | -9.93 | -31.8, 11.9 | 0.4 | -231 | -553, 90.5 | 0.2 |
| PC2 | 14.5 | -4.19, 33.3 | 0.13 | 82.2 | -17.7, 182 | 0.11 |
| PC3 | -19.0 | -39.2, 1.07 | 0.064 | -21.9 | -42.2, -1.55 | 0.035 |
| PC4 | -2.20 | -23.6, 19.2 | 0.8 | -2.32 | -23.7, 19.1 | 0.8 |
| PC5 | -13.8 | -28.5, 0.937 | 0.066 | -13.6 | -28.3, 1.16 | 0.071 |
| CENTER | | | | | | |
| Bronx (reference) | | | | | | |
| Chicago | 0.254 | -0.340, 0.848 | 0.4 | 0.276 | -0.319, 0.872 | 0.4 |

| | | | | | | |
|---|---|---|---|---|---|---|
| Miami | 0.180 | -0.390, 0.750 | 0.5 | 0.206 | -0.366, 0.778 | 0.5 |
| San Diego | 0.000 | -0.720, 0.720 | >0.9 | 0.035 | -0.690, 0.760 | >0.9 |
| AGE | 0.328 | 0.231, 0.424 | <0.001 | 0.329 | 0.232, 0.425 | <0.001 |
| AGE squared | -0.004 | -0.005, -0.003 | <0.001 | -0.004 | -0.005, -0.003 | <0.001 |
| GENDER | | | | | | |
| Female (reference) | | | | | | |
| Male | -1.22 | -1.63, -0.806 | <0.001 | -1.21 | -1.62, -0.805 | <0.001 |
| Type 2 Diabetes History | 1.92 | 1.45, 2.39 | <0.001 | 1.92 | 1.45, 2.39 | <0.001 |
| Sleep Duration (h/day) | -0.314 | -0.445, -0.184 | <0.001 | -0.315 | -0.446, -0.185 | <0.001 |
| CIGARETTE USE | | | | | | |
| Never (reference) | | | | | | |
| Former | 0.736 | 0.288, 1.18 | 0.001 | 0.734 | 0.287, 1.18 | 0.001 |
| Current | -0.876 | -1.36, -0.393 | <0.001 | -0.870 | -1.35, -0.387 | <0.001 |
| Meets 2008 Physical Activity Guidelines | -0.499 | -0.852, -0.147 | 0.006 | -0.495 | -0.848, -0.142 | 0.006 |
| Prevalent CVD | 1.32 | 0.504, 2.14 | 0.002 | 1.32 | 0.501, 2.14 | 0.002 |
| Alcohol Use Level | | | | | | |
| Non-drinker (reference) | | | | | | |

| | | | | | | |
|---|---|---|---|---|---|---|
| Former | -0.430 | -0.808, -0.051 | 0.026 | -0.425 | -0.802, -0.047 | 0.028 |
| Current | 0.415 | -0.360, 1.19 | 0.3 | 0.405 | -0.369, 1.18 | 0.3 |
| Sweetened Beverage Consumption (svgs/day) | 0.162 | -0.077, 0.401 | 0.2 | 0.162 | -0.077, 0.401 | 0.2 |
| Immigrant Generation (1st) | 0.703 | -1.77, 3.17 | 0.6 | 0.758 | -1.70, 3.21 | 0.5 |
| Employment Status | | | | | | |
| Retired and not currently working (reference) | | | | | | |
| Not retired and not currently working | -0.752 | -1.42, -0.085 | 0.027 | -0.755 | -1.42, -0.089 | 0.026 |
| Employed <= 35h/wk | -1.27 | -2.03, -0.510 | 0.001 | -1.28 | -2.03, -0.517 | <0.001 |
| Employed > 35h/wk | -1.26 | -1.94, -0.588 | <0.001 | -1.27 | -1.95, -0.595 | <0.001 |
| Education (Less than HS) | -0.167 | -0.564, 0.230 | 0.4 | -0.157 | -0.554, 0.241 | 0.4 |
| Age at Immigration | | | | | | |
| >= 21 years (reference) | | | | | | |
| US Born | 1.43 | -1.12, 3.98 | 0.3 | 1.38 | -1.15, 3.92 | 0.3 |
| 0-5 years | 2.42 | 1.30,3.54 | <0.001 | 2.42 | 1.31, 3.54 | <0.001 |
| 6-12 years | 1.34 | 0.483, 2.19 | 0.002 | 1.37 | 0.515, 2.22 | 0.002 |

| | | | | | | |
|---|---|---|---|---|---|---|
| 13-20 years | 0.369 | -0.117, 0.854 | 0.14 | 0.368 | -0.117, 0.853 | 0.14 |
| Healthy Diet (Top 40th Percentile Diet Score) | -0.639 | -1.03, -0.246 | 0.001 | -0.644 | -1.04, -0.250 | 0.001 |
| Income | -0.162 | -0.561, 0.237 | 0.4 | -0.166 | -0.565, 0.233 | 0.4 |
| $PGS_{BMI}$ * Healthy Diet | -0.403 | -0.732, -0.075 | 0.016 | -0.398 | -0.725, -0.070 | 0.017 |
| $PGS_{BMI}$ * Age at immigration (Reference = >=21 years) | | | | | | |
| $PGS_{BMI}$ * US Born | 0.522 | -0.120, 1.16 | 0.11 | 0.514 | -0.126, 1.15 | 0.12 |
| $PGS_{BMI}$ * 0-5 years | 0.906 | 0.114, 1.70 | 0.025 | 0.914 | 0.123,1.70 | 0.023 |
| $PGS_{BMI}$ * 6-12 years | 0.187 | -0.731, 1.11 | 0.7 | 0.226 | -0.678, 1.13 | 0.6 |
| $PGS_{BMI}$ * 13-20 years | -0.167 | -0.585, 0.252 | 0.4 | -0.162 | -0.580, 0.256 | 0.4 |
| AME ancestry | NA | NA | NA | 9.22 | -4.13, 22.6 | 0.2 |

Supplemental Table 2. Regression Coefficients for Models Stratified by AME Ancestry Quartiles

| | Quartile 1 | | | Quartile 2 | | | Quartile 3 | | | Quartile 4 | | |
|---|---|---|---|---|---|---|---|---|---|---|---|---|
| **Characteristic** | **Beta** | **95% CI** | **p-value** | **Beta** | **95% CI** | **p-value** | **Beta** | **95% CI** | **p-value** | **Beta** | **95% CI** | **p-value** |
| $PGS_{BMI}$ | 2.23 | 1.32, 3.14 | <0.001 | 1.89 | 1.11, 2.67 | <0.001 | 2.73 | 1.72, 3.74 | <0.001 | 2.40 | 1.27, 3.52 | <0.001 |
| PC1 | -92.6 | -348, 162 | 0.5 | -88.7 | -226, 48.6 | 0.2 | -34.8 | -163, 93.0 | 0.6 | -102 | -193, -11.4 | 0.027 |
| PC2 | 25.6 | -48.4, 99.6 | 0.5 | 14.0 | -44.9, 72.8 | 0.6 | 11.2 | -67.1, 89.5 | 0.8 | -75.3 | -171, 20.6 | 0.12 |
| PC3 | 20.0 | -135, 175 | 0.8 | -41.7 | -118, 34.2 | 0.3 | 84.1 | -12.9, 181 | 0.090 | 41.0 | -67.2, 149 | 0.5 |
| PC4 | 19.1 | -122, 161 | 0.8 | -19.7 | -98.9, 59.5 | 0.6 | -82.7 | -166, 0.762 | 0.052 | -34.6 | -128, 58.4 | 0.5 |
| PC5 | 118 | -6.81, 242 | 0.064 | -56.5 | -139, 26.2 | 0.2 | -48.7 | -84.0, -13.4 | 0.007 | -16.8 | -37.1, 3.46 | 0.10 |
| CENTER | | | | | | | | | | | | |
| Bronx (reference) | | | | | | | | | | | | |
| Chicago | 1.54 | -0.519, 3.59 | 0.14 | 0.061 | -0.781, 0.904 | 0.9 | 0.318 | -1.24, 1.88 | 0.7 | 0.178 | -0.487, 0.842 | 0.6 |
| Miami | 1.14 | 0.378, 1.90 | 0.003 | -0.502 | -1.45, 0.448 | 0.3 | -0.587 | -1.97, 0.793 | 0.4 | -0.229 | -1.16, 0.703 | 0.6 |
| San Diego | -0.229 | -2.23, 1.77 | 0.8 | -0.262 | -1.58, 1.06 | 0.7 | 0.771 | -0.802, 2.34 | 0.3 | -0.377 | -1.13, 0.372 | 0.3 |
| AGE | 0.396 | 0.270, 0.521 | <0.001 | 0.311 | 0.168, 0.455 | <0.001 | 0.215 | 0.085, 0.344 | 0.001 | 0.463 | 0.335, 0.592 | <0.001 |
| AGE squared | -0.004 | -0.006, -0.003 | <0.001 | -0.003 | -0.005, -0.002 | <0.001 | -0.003 | -0.004, -0.001 | <0.001 | -0.005 | -0.006, -0.003 | <0.001 |

| | | | | | | | | | | | | |
|---|---|---|---|---|---|---|---|---|---|---|---|---|
| GENDER | | | | | | | | | | | | |
| Female (reference) | | | | | | | | | | | | |
| Male | -0.893 | -1.45, -0.335 | 0.002 | -1.45 | -2.02, -0.870 | <0.001 | -1.60 | -2.20, -1.00 | <0.001 | -0.98 | -1.52, -0.439 | <0.001 |
| Type 2 Diabetes History | 1.77 | 1.08, 2.47 | <0.001 | 2.60 | 1.84, 3.36 | <0.001 | 1.90 | 1.19, 2.61 | <0.001 | 1.61 | 0.98, 2.25 | <0.001 |
| Sleep Duration (h/day) | -0.310 | -0.481, -0.139 | <0.001 | -0.416 | -0.596, -0.235 | <0.001 | -0.125 | -0.325, 0.076 | 0.2 | -0.182 | -0.354, -0.009 | 0.039 |
| CIGARETTE USE | | | | | | | | | | | | |
| Never (reference) | | | | | | | | | | | | |
| Former | 0.001 | -0.664, 0.666 | >0.9 | 0.907 | 0.213, 1.60 | 0.011 | 1.55 | 0.900, 2.20 | <0.001 | 0.802 | 0.213, 1.39 | 0.008 |
| Current | -1.83 | -2.46, -1.19 | <0.001 | -0.932 | -1.57, -0.296 | 0.004 | -0.528 | -1.22, 0.161 | 0.13 | 0.243 | -0.408, 0.894 | 0.5 |
| Meets 2008 Physical Activity Guidelines | -1.13 | -1.64, -0.625 | <0.001 | -0.602 | -1.18, -0.025 | 0.041 | -0.237 | -0.791, 0.318 | 0.4 | -0.103 | -0.586, 0.381 | 0.7 |
| Prevalent CVD | 0.385 | -0.527, 1.30 | 0.4 | 1.64 | 0.669, 2.62 | <0.001 | 2.57 | 1.40, 3.74 | <0.001 | 1.10 | 0.097, 2.11 | 0.032 |
| Alcohol Use Level | | | | | | | | | | | | |
| Non-drinker (reference) | | | | | | | | | | | | |
| Former | -0.328 | -0.845, 0.188 | 0.2 | -0.486 | -1.05, 0.077 | 0.091 | -1.03 | -1.56, -0.496 | <0.001 | 0.047 | -0.411, 0.505 | 0.8 |
| Current | 1.00 | -0.061, 2.06 | 0.065 | -0.464 | -1.54, 0.609 | 0.4 | 0.927 | -0.126, 1.98 | 0.085 | 0.008 | -1.03, 1.05 | >0.9 |

| | | | | | | | | | | | | |
|---|---|---|---|---|---|---|---|---|---|---|---|---|
| Sweetened Beverage Consumption (svgs/day) | -0.431 | -0.761, -0.102 | 0.010 | 0.172 | -0.108, 0.452 | 0.2 | 0.489 | 0.201, 0.778 | <0.001 | 0.120 | -0.127, 0.366 | 0.3 |
| Immigrant Generation (1st) | 7.64 | 4.65, 10.6 | <0.001 | -0.331 | -2.28, 1.62 | 0.7 | 0.105 | -1.40, 1.61 | 0.9 | -0.619 | -3.99, 2.75 | 0.7 |
| Employment Status | | | | | | | | | | | | |
| Retired and not currently working (reference) | | | | | | | | | | | | |
| Not retired and not currently working | -1.07 | -2.02, -0.122 | 0.027 | -0.587 | -1.61, 0.436 | 0.3 | -1.49 | -2.88, -0.095 | 0.036 | 0.839 | -0.528, 2.21 | 0.2 |
| Employed <= 35h/wk | -1.32 | -2.42, -0.221 | 0.019 | -0.598 | -1.77, 0.570 | 0.3 | -2.24 | -3.69, -0.794 | 0.002 | -0.115 | -1.52, 1.29 | 0.9 |
| Employed > 35h/wk | -1.25 | -2.25, -0.246 | 0.015 | -1.13 | -2.21, -0.057 | 0.039 | -2.00 | -3.41, -0.585 | 0.006 | 0.061 | -1.33, 1.45 | >0.9 |
| Education (Less than HS) | 0.104 | -0.494, 0.702 | 0.7 | 0.668 | 0.045, 1.29 | 0.036 | -0.303 | -0.850, 0.244 | 0.3 | -0.631 | -1.10, -0.159 | 0.009 |
| Age at Immigration | | | | | | | | | | | | |
| >= 21 years (reference) | REF | REF | REF | REF | REF | REF | REF | REF | REF | REF | REF | REF |
| US Born | -5.23 | -8.25,-2.20 | <0.001 | 1.69 | -0.323, 3.71 | 0.10 | 2.36 | 0,714, 4.00 | 0.005 | 3.57 | 0.089, 7.04 | 0.045 |
| 0-5 years | 2.89 | 1.27,4.51 | <0.001 | 1.96 | 0.911, 3.01 | <0.001 | 0.729 | -0.459, 1.92 | 0.2 | 3.57 | 2.08,5.07 | <0.001 |
| 6-12 years | 2.90 | 1.68,4.12 | <0.001 | 0.490 | -0.717, 1.70 | 0.4 | 0.884 | -0.374, 2.14 | 0.2 | 1.71 | 0.543, 2.87 | 0.004 |

| | | | | | | | | | | | | |
|---|---|---|---|---|---|---|---|---|---|---|---|---|
| 13-20 years | 0.137 | -0.673, 0.947 | 0.7 | -0.334 | -1.22,0.553 | 0.5 | 0.052 | -0.631, 0.734 | 0.9 | 1.53 | 0.963, 2.10 | <0.001 |
| Healthy Diet (Top 40th Percentile Diet Score) | -0.137 | -0.671, 0.397 | 0.6 | -0.807 | -1.44, -0.171 | 0.013 | -1.40 | -1.96, -0.840 | <0.001 | -0.079 | -0.578, 0.421 | 0.8 |
| Income | -0.057 | -0.608, 0.493 | 0.8 | -0.309 | -0.930, 0.311 | 0.3 | 0.103 | -0.438, 0.645 | 0.7 | -0.602 | -1.10, -0.101 | 0.019 |
| $PGS_{BMI}$ * Healthy Diet | -0.440 | -0.921, 0.042 | 0.074 | -0.226 | -0.745, 0.294 | 0.4 | -0.367 | -0.891, 0.158 | 0.2 | -0.536 | -1.04, -0.030 | 0.038 |
| $PGS_{BMI}$ * Age at immigration (Reference = >=21 years) | | | | | | | | | | | | |
| $PGS_{BMI}$ * US Born | 0.027 | -0.689, 0.743 | >0.9 | 0.566 | -0.030, 1.16 | 0.063 | 0.788 | 0.183, 1.29 | 0.011 | 0.179 | -0.746, 1.10 | 0.7 |
| $PGS_{BMI}$ * 0-5 years | 0.686 | -0.567, 1.94 | 0.3 | 1.48 | 0.606, 2.35 | <0.001 | 1.03 | -0.281, 2.34 | 0.12 | 0.376 | -0.954, 1.71 | 0.6 |
| $PGS_{BMI}$ * 6-12 years | 0.491 | -0.772, 1.75 | 0.4 | 0.725 | -0.510, 1.96 | 0.3 | 0.460 | -0.759, 1.68 | 0.5 | -0.259 | -1.59,1.08 | 0.7 |
| $PGS_{BMI}$ * 13-20 years | 0.288 | -0.485, 1.06 | 0.5 | 0.142 | -0.653, 0.937 | 0.7 | 0.153 | -0.550, 0.857 | 0.7 | -0.907 | -1.49,-0.323 | 0.002 |

Supplemental Tables 3.1 - 3.6. Regression Coefficients for Models Stratified by Ancestry Tertiles & Background Group

*Tertiles with extremely low sample sizes for which regressions could not be run are not shown in the below tables.*

3.1. Central American

| | Tertile 2 | | | Tertile 3 | | |
|---|---|---|---|---|---|---|
| **Characteristic** | **Beta** | **95% CI** | **p-value** | **Beta** | **95% CI** | **p-value** |
| $PGS_{BMI}$ | 1.19 | -0.623,3.00 | 0.2 | 2.70 | 1.03,4.37 | 0.002 |
| PC1 | -198 | -487, 91.2 | 0.2 | 78.9 | -92.0, 250 | 0.4 |
| PC2 | 48.7 | -80.6, 178 | 0.5 | 166 | 19.6, 313 | 0.027 |
| PC3 | 340 | 69.7, 610 | 0.014 | -60.3 | -318, 198 | 0.6 |
| PC4 | -288 | -448, -129 | <0.001 | 52.7 | -160, 266 | 0.6 |
| PC5 | -63.4 | -209, 81.8 | 0.4 | -28.2 | -135, 78.7 | 0.6 |
| CENTER | | | | | | |
| Bronx (reference) | | | | | | |
| Chicago | -1.07 | -3.55, 1.40 | 0.4 | -0.040 | -1.74, 1.66 | >0.9 |
| Miami | -2.29 | -4.38, -0.205 | 0.032 | 0.908 | -0.689, 2.50 | 0.3 |
| San Diego | -2.87 | -5.84, 0.095 | 0.059 | -1.18 | -3.49, 1.12 | 0.3 |
| AGE | 0.596 | 0.308, 0.885 | <0.001 | 0.636 | 0.357, 0.915 | <0.001 |
| AGE squared | -0.006 | -0.009, -0.003 | <0.001 | -0.007 | -0.010, -0.004 | <0.001 |
| GENDER | | | | | | |
| Female (reference) | | | | | | |
| Male | -1.90 | -3.19, -0.611 | 0.004 | -1.03 | -2.20, 0.134 | 0.083 |
| Type 2 Diabetes History | 2.38 | 0.849, 3.92 | 0.003 | 1.04 | -0.439, 2.52 | 0.2 |
| Sleep Duration (h/day) | -0.273 | -0.685, 0.139 | 0.2 | -0.331 | -0.705, 0.043 | 0.084 |
| CIGARETTE USE | | | | | | |
| Never (reference) | | | | | | |
| Former | 1.45 | 0.081, 2.82 | 0.039 | 0.99 | -0.340, 2.32 | 0.15 |
| Current | -0.567 | -2.22, 1.08 | 0.5 | -0.535 | -2.05, 0.98 | 0.5 |
| Meets 2008 Physical Activity Guidelines | -0.662 | -1.84, 0.511 | 0.3 | 0.483 | -0.561, 1.53 | 0.4 |
| Prevalent CVD | -0.371 | -3.31, 2.56 | 0.8 | 1.14 | -0.95, 3.23 | 0.3 |

| | | | | | | |
|---|---|---|---|---|---|---|
| Alcohol Use Level | | | | | | |
| Non-drinker (reference) | | | | | | |
| Former | -0.213 | -1.40, 0.97 | 0.7 | 0.636 | -0.383, 1.65 | 0.2 |
| Current | -0.582 | -2.89, 1.72 | 0.6 | -1.56 | -3.58, 0.471 | 0.13 |
| Sweetened Beverage Consumption (svgs/day) | 0.98 | 0.344, 1.62 | 0.003 | -0.900 | -1.49, -0.311 | 0.003 |
| Immigrant Generation (1st) | 0.200 | -2.38, 2.79 | 0.9 | -0.886 | -3.92, 2.15 | 0.6 |
| Employment Status | | | | | | |
| Retired and not currently working (reference) | | | | | | |
| Not retired and not currently working | 0.007 | -3.21, 3.23 | >0.9 | -2.57 | -5.51, 0.365 | 0.087 |
| Employed <= 35h/wk | 0.155 | -3.14, 3.45 | >0.9 | -3.93 | -6.93, -0.941 | 0.010 |
| Employed > 35h/wk | 0.377 | -2.95, 3.71 | 0.8 | -3.86 | -6.81, -0.906 | 0.011 |
| Education (Less than HS) | -0.299 | -1.45, 0.856 | 0.6 | -1.01 | -1.99, -0.023 | 0.046 |
| Age at Immigration | | | | | | |
| >= 21 years (reference) | REF | REF | REF | REF | REF | REF |
| US Born | N/A | N/A | N/A | N/A | N/A | N/A |
| 0-5 years | 0.890 | -4.25, 6.03 | 0.7 | 6.15 | 1.37, 10.9 | 0.012 |
| 6-12 years | 2.69 | -0.495, 5.88 | 0.10 | 1.71 | -0.884, 4.31 | 0.2 |
| 13-20 years | 0.838 | -0.702, 2.38 | 0.3 | 2.35 | 0.96, 3.73 | <0.001 |
| Healthy Diet (Top 40th Percentile Diet Score) | 0.577 | -0.557, 1.71 | 0.3 | -0.182 | -1.22, 0.859 | 0.7 |
| Income | -0.592 | -1.88, 0.695 | 0.4 | -0.108 | -1.30, 1.08 | 0.9 |
| $PGS_{BMI}$ * Healthy Diet | 0.447 | -0.656, 1.55 | 0.4 | -1.03 | -2.10, 0.038 | 0.059 |
| $PGS_{BMI}$ * Age at immigration (Reference = >=21 years) | | | | | | |
| $PGS_{BMI}$ * US Born | 0.210 | -1.80, 2.22 | 0.8 | 2.40 | -0.497, 5.30 | 0.11 |
| $PGS_{BMI}$ * 0-5 years | -1.48 | -4.93, 1.97 | 0.4 | -3.57 | -9.29, 2.14 | 0.2 |
| $PGS_{BMI}$ * 6-12 years | -0.557 | -4.55,3.43 | 0.8 | -0.408 | -2.95,2.13 | 0.8 |
| $PGS_{BMI}$ * 13-20 years | 1.53 | -0.303,3.36 | 0.10 | -0.713 | -2.06,0.635 | 0.3 |

3.2. Cuban

| | Tertile 1 | | | Tertile 2 | | |
|---|---|---|---|---|---|---|
| **Characteristic** | **Beta** | **95% CI** | **p-value** | **Beta** | **95% CI** | **p-value** |
| $PGS_{BMI}$ | 2.06 | 1.24,2.89 | <0.001 | 3.50 | 0.641,6.35 | 0.019 |
| PC1 | -120 | -385, 145 | 0.4 | -530 | -1,494, 434 | 0.3 |
| PC2 | 43.5 | -27.9, 115 | 0.2 | 150 | -164, 464 | 0.4 |
| PC3 | 91.8 | -127, 310 | 0.4 | 439 | -116, 993 | 0.13 |
| PC4 | -51.8 | -253, 150 | 0.6 | -224 | -691, 242 | 0.3 |
| PC5 | 186 | 31.1, 340 | 0.019 | 73.0 | -308, 454 | 0.7 |
| CENTER | | | | | | |
| Bronx (reference) | | | | | | |
| Chicago | 1.16 | -2.09, 4.42 | 0.5 | -5.07 | -13.3, 3.15 | 0.2 |
| Miami | 1.28 | -0.102, 2.67 | 0.070 | -3.50 | -16.9, 9.89 | 0.6 |
| San Diego | 0.166 | -3.74, 4.07 | >0.9 | -0.327 | -10.8, 10.2 | >0.9 |
| AGE | 0.483 | 0.329, 0.637 | <0.001 | 0.879 | 0.201, 1.56 | 0.013 |
| AGE squared | -0.005 | -0.007, -0.003 | <0.001 | -0.009 | -0.016, -0.001 | 0.022 |
| GENDER | | | | | | |
| Female (reference) | | | | | | |
| Male | -0.614 | -1.29, 0.061 | 0.075 | 0.107 | -2.85, 3.07 | >0.9 |
| Type 2 Diabetes History | 2.08 | 1.25, 2.91 | <0.001 | 1.26 | -1.99, 4.52 | 0.4 |
| Sleep Duration (h/day) | -0.234 | -0.453, -0.015 | 0.037 | 0.016 | -0.820, 0.852 | >0.9 |
| CIGARETTE USE | | | | | | |
| Never (reference) | | | | | | |
| Former | -0.554 | -1.33, 0.222 | 0.2 | 1.21 | -1.40, 3.82 | 0.4 |
| Current | -2.27 | -2.99, -1.55 | <0.001 | -0.884 | -3.54, 1.77 | 0.5 |
| Meets 2008 Physical Activity Guidelines | -1.33 | -1.93, -0.723 | <0.001 | -0.636 | -2.61, 1.34 | 0.5 |
| Prevalent CVD | -0.031 | -1.09, 1.03 | >0.9 | -1.58 | -5.21, 2.05 | 0.4 |
| Alcohol Use Level | | | | | | |
| Non-drinker (reference) | | | | | | |
| Former | -0.056 | -0.675, 0.564 | 0.9 | 0.561 | -1.92, 3.04 | 0.7 |

| | | | | | | |
|---|---|---|---|---|---|---|
| Current | 1.60 | 0.344, 2.85 | 0.013 | -3.97 | -8.76, 0.822 | 0.11 |
| Sweetened Beverage Consumption (svgs/day) | -0.404 | -0.808, 0.000 | 0.050 | 0.674 | -0.651, 2.00 | 0.3 |
| Immigrant Generation (1st) | 0.417 | -6.39, 7.22 | >0.9 | -1.36 | -19.8, 17.1 | 0.9 |
| Employment Status | | | | | | |
| Retired and not currently working (reference) | | | | | | |
| Not retired and not currently working | -0.788 | -1.93, 0.350 | 0.2 | -0.899 | -6.15, 4.35 | 0.7 |
| Employed <= 35h/wk | -1.56 | -2.91, -0.204 | 0.024 | -0.96 | -7.09, 5.17 | 0.8 |
| Employed > 35h/wk | -1.13 | -2.33, 0.069 | 0.065 | -0.609 | -6.47, 5.25 | 0.8 |
| Education (Less than HS) | 0.534 | -0.230, 1.30 | 0.2 | -3.72 | -6.58, -0.853 | 0.013 |
| Age at Immigration | | | | | | |
| >= 21 years (reference) | REF | REF | REF | REF | REF | REF |
| US Born | 2.67 | -4.22,9.56 | 0.4 | 1.57 | -13.3.16.4 | 0.8 |
| 0-5 years | 0.651 | -1.44,2.74 | 0.5 | N/A | N/A | N/A |
| 6-12 years | 1.65 | 0.013,3.29 | 0.048 | 2.83 | -18.1,23.7 | 0.8 |
| 13-20 years | -0.073 | -1.16,1.02 | 0.9 | 1.33 | -3.74,6.40 | 0.6 |
| Healthy Diet (Top 40th Percentile Diet Score) | 0.268 | -0.348, 0.884 | 0.4 | 1.58 | -0.478,3.63 | 0.14 |
| Income | -0.097 | -0.764, 0.570 | 0.8 | -0.574 | -2.89, 1.74 | 0.6 |
| $PGS_{BMI}$ * Healthy Diet | -0.292 | -0.834, 0.251 | 0.3 | -2.32 | -4.22, -0.424 | 0.019 |
| $PGS_{BMI}$ * Age at immigration (Reference = >=21 years) | REF | REF | REF | REF | REF | REF |
| $PGS_{BMI}$ * US Born | 0.378 | -0.564, 1.32 | 0.4 | 3.97 | -0.721, 8.66 | 0.10 |
| $PGS_{BMI}$ * 0-5 years | 2.20 | 0.427, 3.98 | 0.015 | N/A | N/A | N/A |
| $PGS_{BMI}$ * 6-12 years | -1.28 | -2.90,0.336 | 0.12 | 6.16 | -13.5,25.8 | 0.5 |
| $PGS_{BMI}$ * 13-20 years | 0.383 | -0.605,1.37 | 0.4 | 2.70 | -3.46,8.85 | 0.4 |

3.3. Dominican

| | Tertile 1 | | |
|---|---|---|---|
| **Characteristic** | **Beta** | **95% CI** | **p-value** |
| $PGS_{BMI}$ | 2.15 | 0.679,3.63 | 0.004 |
| PC1 | 865 | 269, 1,462 | 0.005 |
| PC2 | -390 | -594, -185 | <0.001 |
| PC3 | 11.9 | -288, 312 | >0.9 |
| PC4 | 103 | -148, 354 | 0.4 |
| PC5 | 30.7 | -150, 211 | 0.7 |
| CENTER | | | |
| Bronx (reference) | | | |
| Chicago | -0.226 | -4.26, 3.81 | >0.9 |
| Miami | 3.03 | 1.15, 4.91 | 0.002 |
| San Diego | -0.879 | -5.14, 3.38 | 0.7 |
| AGE | 0.314 | 0.081, 0.547 | 0.008 |
| AGE squared | -0.004 | -0.007, -0.001 | 0.002 |
| GENDER | | | |
| Female (reference) | | | |
| Male | -1.28 | -2.22, -0.343 | 0.008 |
| Type 2 Diabetes History | 1.77 | 0.566, 2.97 | 0.004 |
| Sleep Duration (h/day) | -0.364 | -0.620, -0.109 | 0.005 |
| CIGARETTE USE | | | |
| Never (reference) | | | |
| Former | 1.55 | 0.300, 2.81 | 0.015 |
| Current | 0.516 | -0.905, 1.94 | 0.5 |
| Meets 2008 Physical Activity Guidelines | -0.460 | -1.30, 0.376 | 0.3 |
| Prevalent CVD | 2.97 | 1.26, 4.67 | <0.001 |
| Alcohol Use Level | | | |
| Non-drinker (reference) | | | |
| Former | -0.783 | -1.61, 0.047 | 0.065 |
| Current | -0.896 | -2.74, 0.948 | 0.3 |

| | | | |
|---|---|---|---|
| Sweetened Beverage Consumption (svgs/day) | -0.032 | -0.626, 0.562 | >0.9 |
| Immigrant Generation (1st) | 0.726 | -13.9, 15.3 | >0.9 |
| Employment Status | | | |
| Retired and not currently working (reference) | | | |
| Not retired and not currently working | -2.45 | -4.20, -0.700 | 0.006 |
| Employed <= 35h/wk | -2.14 | -4.10, -0.181 | 0.033 |
| Employed > 35h/wk | -2.22 | -4.01, -0.433 | 0.015 |
| Education (Less than HS) | -0.212 | -1.08, 0.659 | 0.6 |
| Age at Immigration | | | |
| >= 21 years (reference) | | | |
| US Born | 0.789 | -13.9,15.5 | >0.9 |
| 0-5 years | 2.18 | -0.302,4.66 | 0.086 |
| 6-12 years | 1.92 | 0.070,3.77 | 0.042 |
| 13-20 years | -0.274 | -1.44,0.897 | 0.6 |
| Healthy Diet (Top 40th Percentile Diet Score) | -0.526 | -1.53, 0.477 | 0.3 |
| Income | 0.249 | -0.682, 1.18 | 0.6 |
| $PGS_{BMI}$ * Healthy Diet | -0.677 | -1.75, 0.397 | 0.2 |
| $PGS_{BMI}$ * Age at immigration (Reference = >=21 years) | | | |
| $PGS_{BMI}$ * US Born | 0.274 | -1.56,2.11 | 0.8 |
| $PGS_{BMI}$ * 0-5 years | -0.492 | -2.17,1.18 | 0.6 |
| $PGS_{BMI}$ * 6-12 years | 4.38 | 2.14,6.62 | <0.001 |
| $PGS_{BMI}$ * 13-20 years | -0.622 | -1.77,0.530 | 0.3 |

3.4. Mexican

| | Tertile 2 | | | Tertile 3 | | |
|---|---|---|---|---|---|---|
| **Characteristic** | **Beta** | **95% CI** | **p-value** | **Beta** | **95% CI** | **p-value** |
| $PGS_{BMI}$ | 3.25 | 1.72,4.77 | <0.001 | 1.21 | 0.190,2.22 | 0.020 |
| PC1 | 118 | -183, 420 | 0.4 | -174 | -314, -33.8 | 0.015 |
| PC2 | -144 | -338, 50.7 | 0.15 | -27.5 | -175, 120 | 0.7 |
| PC3 | 304 | 85.6, 523 | 0.006 | 107 | -28.1, 243 | 0.12 |
| PC4 | 277 | 88.5, 466 | 0.004 | -232 | -358, -105 | <0.001 |
| PC5 | 46.3 | -80.8, 173 | 0.5 | 56.4 | -23.0, 136 | 0.2 |
| CENTER | | | | | | |
| Bronx (reference) | | | | | | |
| Chicago | 4.76 | -9.09, 18.6 | 0.5 | -0.216 | -1.04, 0.603 | 0.6 |
| Miami | 2.62 | -11.5, 16.7 | 0.7 | -2.66 | -4.58, -0.734 | 0.007 |
| San Diego | 4.96 | -8.87, 18.8 | 0.5 | -0.575 | -1.45, 0.302 | 0.2 |
| AGE | 0.135 | -0.037, 0.308 | 0.12 | 0.290 | 0.156, 0.423 | <0.001 |
| AGE squared | -0.002 | -0.004, 0.000 | 0.087 | -0.003 | -0.005, -0.002 | <0.001 |
| GENDER | | | | | | |
| Female (reference) | | | | | | |
| Male | -1.68 | -2.46, -0.903 | <0.001 | -1.28 | -1.86, -0.706 | <0.001 |
| Type 2 Diabetes History | 2.31 | 1.35, 3.26 | <0.001 | 1.60 | 0.940, 2.26 | <0.001 |
| Sleep Duration (h/day) | -0.444 | -0.724, -0.165 | 0.002 | -0.030 | -0.215, 0.155 | 0.8 |
| CIGARETTE USE | | | | | | |
| Never (reference) | | | | | | |
| Former | 1.65 | 0.838, 2.47 | <0.001 | 0.837 | 0.194, 1.48 | 0.011 |
| Current | -0.542 | -1.45, 0.370 | 0.2 | 0.179 | -0.483, 0.842 | 0.6 |
| Meets 2008 Physical Activity Guidelines | 0.042 | -0.683, 0.767 | >0.9 | -0.144 | -0.665, 0.378 | 0.6 |
| Prevalent CVD | 0.731 | -0.654, 2.12 | 0.3 | 2.46 | 1.30, 3.62 | <0.001 |
| Alcohol Use Level | | | | | | |
| Non-drinker (reference) | | | | | | |
| Former | -1.34 | -2.03, -0.655 | <0.001 | -0.220 | -0.716, 0.275 | 0.4 |
| Current | 0.447 | -0.797, 1.69 | 0.5 | 0.876 | -0.192, 1.94 | 0.11 |

| | | | | | | |
|---|---|---|---|---|---|---|
| Sweetened Beverage Consumption (svgs/day) | 0.581 | 0.222, 0.939 | 0.002 | 0.192 | -0.077, 0.460 | 0.2 |
| Immigrant Generation (1st) | -0.742 | -2.22, 0.738 | 0.3 | 1.64 | -0.95, 4.24 | 0.2 |
| Employment Status | | | | | | |
| Retired and not currently working (reference) | | | | | | |
| Not retired and not currently working | -2.18 | -3.79, -0.572 | 0.008 | 1.58 | 0.042, 3.12 | 0.044 |
| Employed <= 35h/wk | -2.34 | -3.98, -0.690 | 0.006 | 0.438 | -1.14, 2.01 | 0.6 |
| Employed > 35h/wk | -2.17 | -3.77, -0.568 | 0.008 | 0.782 | -0.782, 2.35 | 0.3 |
| Education (Less than HS) | 0.259 | -0.540, 1.06 | 0.5 | -0.532 | -1.03, -0.034 | 0.036 |
| Age at Immigration | | | | | | |
| >= 21 years (reference) | | | | | | |
| US Born | 2.75 | 1.10, 4.40 | 0.001 | 0.874 | -1.84,3.59 | 0.5 |
| 0-5 years | -1.02 | -2.40, 0.359 | 0.15 | 3.74 | 2.39,5.10 | <0.001 |
| 6-12 years | -0.265 | -1.91,1.37 | 0.8 | 1.40 | 0.182,2.61 | 0.024 |
| 13-20 years | -1.09 | -2.00,-0.175 | 0.020 | 0.825 | 0.232,1.42 | 0.006 |
| Healthy Diet (Top 40th Percentile Diet Score) | -1.17 | -1.97, -0.371 | 0.004 | -0.364 | -0.899, 0.171 | 0.2 |
| Income | -0.205 | -0.887, 0.477 | 0.6 | -0.382 | -0.911, 0.146 | 0.2 |
| $PGS_{BMI}$ * Healthy Diet | -1.18 | -1.95, -0.412 | 0.003 | -0.053 | -0.598, 0.492 | 0.8 |
| $PGS_{BMI}$ * Age at immigration (Reference = >=21 years) | | | | | | |
| $PGS_{BMI}$ * US Born | 1.33 | 0.590,2.07 | <0.001 | -0.043 | -0.809,0.723 | >0.9 |
| $PGS_{BMI}$ * 0-5 years | 1.17 | -0.371,2.70 | 0.14 | 0.752 | -0.451,1.95 | 0.2 |
| $PGS_{BMI}$ * 6-12 years | -0.207 | -1.73,1.32 | 0.8 | 0.572 | -0.780,1.93 | 0.4 |
| $PGS_{BMI}$ * 13-20 years | 0.333 | -0.623,1.29 | 0.5 | -0.545 | -1.16,0.066 | 0.081 |

3.5. Puerto Rican

| | Tertile 1 | | | Tertile 2 | | |
|---|---|---|---|---|---|---|
| **Characteristic** | **Beta** | **95% CI** | **p-value** | **Beta** | **95% CI** | **p-value** |
| $PGS_{BMI}$ | 0.683 | -1.43,2.79 | 0.5 | 2.15 | 0.473,3.83 | 0.012 |
| PC1 | -58.7 | -941, 824 | 0.9 | -348 | -965, 269 | 0.3 |
| PC2 | 49.3 | -220, 319 | 0.7 | 159 | -31.2, 349 | 0.10 |
| PC3 | -89.4 | -445, 266 | 0.6 | 143 | -145, 431 | 0.3 |
| PC4 | 39.0 | -296, 374 | 0.8 | 340 | 84.5, 595 | 0.009 |
| PC5 | -75.5 | -337, 186 | 0.6 | -327 | -519, -135 | <0.001 |
| CENTER | | | | | | |
| Bronx (reference) | | | | | | |
| Chicago | 1.81 | 0.097, 3.52 | 0.039 | -0.252 | -1.42, 0.917 | 0.7 |
| Miami | -0.504 | -2.88, 1.87 | 0.7 | -0.806 | -2.88, 1.27 | 0.4 |
| San Diego | -0.536 | -3.35, 2.27 | 0.7 | 3.52 | 0.187, 6.85 | 0.039 |
| AGE | 0.785 | 0.486, 1.08 | <0.001 | 0.084 | -0.155, 0.323 | 0.5 |
| AGE squared | -0.009 | -0.012, -0.005 | <0.001 | -0.001 | -0.004, 0.001 | 0.3 |
| GENDER | | | | | | |
| Female (reference) | | | | | | |
| Male | -3.61 | -4.82, -2.40 | <0.001 | -1.64 | -2.67, -0.607 | 0.002 |
| Type 2 Diabetes History | 2.55 | 0.98, 4.12 | 0.002 | 2.90 | 1.73, 4.07 | <0.001 |
| Sleep Duration (h/day) | -0.584 | -0.930, -0.238 | 0.001 | -0.349 | -0.650, -0.047 | 0.024 |
| CIGARETTE USE | | | | | | |
| Never (reference) | | | | | | |
| Former | 0.850 | -0.733, 2.43 | 0.3 | 0.772 | -0.516, 2.06 | 0.2 |
| Current | -1.58 | -2.95, -0.215 | 0.024 | -1.25 | -2.30, -0.197 | 0.020 |
| Meets 2008 Physical Activity Guidelines | -0.002 | -1.29, 1.29 | >0.9 | -0.842 | -1.87, 0.186 | 0.11 |
| Prevalent CVD | 3.17 | 1.06, 5.29 | 0.003 | 1.71 | 0.132, 3.30 | 0.034 |
| Alcohol Use Level | | | | | | |
| Non-drinker (reference) | | | | | | |
| Former | -0.338 | -1.56, 0.884 | 0.6 | -0.640 | -1.65, 0.372 | 0.2 |

| | | | | | | |
|---|---|---|---|---|---|---|
| Current | -0.271 | -3.42, 2.87 | 0.9 | -0.462 | -2.30, 1.38 | 0.6 |
| Sweetened Beverage Consumption (svgs/day) | 0.345 | -0.236, 0.926 | 0.2 | 0.228 | -0.278, 0.735 | 0.4 |
| Immigrant Generation (1st) | 1.72 | -1.91, 5.35 | 0.4 | 2.09 | -2.04, 6.23 | 0.3 |
| Employment Status | | | | | | |
| Retired and not currently working (reference) | | | | | | |
| Not retired and not currently working | -3.28 | -5.30, -1.26 | 0.002 | 0.222 | -1.42, 1.86 | 0.8 |
| Employed <= 35h/wk | -1.65 | -3.89, 0.580 | 0.15 | 0.198 | -1.97, 2.37 | 0.9 |
| Employed > 35h/wk | -3.43 | -5.62, -1.24 | 0.002 | -1.14 | -2.99, 0.698 | 0.2 |
| Education (Less than HS) | -1.24 | -2.60, 0.114 | 0.073 | 1.59 | 0.577, 2.59 | 0.002 |
| Age at Immigration | | | | | | |
| >= 21 years (reference) | | | | | | |
| US Born | -0.724 | -4.42,2.97 | 0.7 | -0.75 | -4.92,3.42 | 0.7 |
| 0-5 years | 0.752 | -1.63,3.13 | 0.5 | 3.49 | 1.86,5.13 | <0.001 |
| 6-12 years | 2.38 | -0.200,4.96 | 0.071 | 0.442 | -1.53,2.41 | 0.7 |
| 13-20 years | -2.30 | -4.31,-0.302 | 0.025 | 0.621 | -0.857,2.10 | 0.4 |
| Healthy Diet (Top 40th Percentile Diet Score) | -0.230 | -1.81, 1.35 | 0.8 | -1.39 | -2.64, -0.138 | 0.030 |
| Income | 0.541 | -0.940, 2.02 | 0.5 | 0.328 | -0.812, 1.47 | 0.6 |
| $PGS_{BMI}$ * Healthy Diet | 0.285 | -1.14, 1.71 | 0.7 | -0.161 | -1.28, 0.96 | 0.8 |
| $PGS_{BMI}$ * Age at immigration (Reference = >=21 years) | | | | | | |
| $PGS_{BMI}$ * US Born | 0.648 | -0.7,2.00 | 0.3 | -0.264 | -1.37,0.845 | 0.6 |
| $PGS_{BMI}$ * 0-5 years | -0.067 | -2.03,1.89 | >0.9 | 1.09 | -0.336,2.51 | 0.13 |
| $PGS_{BMI}$ * 6-12 years | 2.02 | -0.874,4.92 | 0.2 | -0.180 | -2.21,1.85 | 0.9 |
| $PGS_{BMI}$ * 13-20 years | 0.878 | -1.29,3.04 | 0.4 | -0.502 | -1.84,0.838 | 0.5 |

3.6. South American

| | Tertile 2 | | | Tertile 3 | | |
|---|---|---|---|---|---|---|
| **Characteristic** | **Beta** | **95% CI** | **p-value** | **Beta** | **95% CI** | **p-value** |
| $PGS_{BMI}$ | 2.35 | 0.120,4.57 | 0.040 | 0.848 | -1.21,2.90 | 0.4 |
| PC1 | 43.3 | -345, 432 | 0.8 | -557 | -905, -209 | 0.002 |
| PC2 | 18.7 | -123, 161 | 0.8 | -274 | -422, -126 | <0.001 |
| PC3 | -22.4 | -334, 289 | 0.9 | 37.0 | -270, 344 | 0.8 |
| PC4 | -53.6 | -251, 144 | 0.6 | 382 | 149, 614 | 0.001 |
| PC5 | -46.2 | -126, 33.4 | 0.3 | 28.5 | -27.5, 84.5 | 0.3 |
| CENTER | | | | | | |
| Bronx (reference) | | | | | | |
| Chicago | 1.14 | -1.71, 3.98 | 0.4 | 0.067 | -1.55, 1.69 | >0.9 |
| Miami | -0.021 | -2.29, 2.25 | >0.9 | 0.545 | -1.02, 2.11 | 0.5 |
| San Diego | -1.70 | -5.30, 1.90 | 0.4 | -0.039 | -3.12, 3.04 | >0.9 |
| AGE | 0.279 | -0.147, 0.705 | 0.2 | 0.337 | 0.018, 0.656 | 0.040 |
| AGE squared | -0.003 | -0.007, 0.002 | 0.3 | -0.003 | -0.007, 0.000 | 0.054 |
| GENDER | | | | | | |
| Female (reference) | | | | | | |
| Male | 0.762 | -0.798, 2.32 | 0.3 | 0.672 | -0.688, 2.03 | 0.3 |
| Type 2 Diabetes History | 1.80 | -0.215, 3.81 | 0.082 | 2.72 | 0.756, 4.68 | 0.007 |
| Sleep Duration (h/day) | -0.341 | -0.870, 0.188 | 0.2 | -0.448 | -0.835, -0.061 | 0.024 |
| CIGARETTE USE | | | | | | |
| Never (reference) | | | | | | |
| Former | 1.68 | 0.047, 3.31 | 0.045 | 0.667 | -0.585, 1.92 | 0.3 |
| Current | 0.156 | -1.80, 2.12 | 0.9 | 1.77 | -0.230, 3.77 | 0.084 |
| Meets 2008 Physical Activity Guidelines | -2.08 | -3.61, -0.552 | 0.008 | -0.568 | -1.74, 0.604 | 0.3 |
| Prevalent CVD | 0.231 | -2.52, 2.98 | 0.9 | 2.71 | 0.351, 5.07 | 0.025 |
| Alcohol Use Level | | | | | | |
| Non-drinker (reference) | | | | | | |
| Former | 0.176 | -1.24, 1.59 | 0.8 | 0.195 | -0.868, 1.26 | 0.7 |

| | | | | | | |
|---|---|---|---|---|---|---|
| Current | -1.25 | -4.22, 1.72 | 0.4 | -0.723 | -7.19, 5.74 | 0.8 |
| Sweetened Beverage Consumption (svgs/day) | -0.068 | -0.780, 0.645 | 0.9 | -0.103 | -0.701, 0.496 | 0.7 |
| Immigrant Generation (1st) | 0.676 | -1.98, 3.33 | 0.6 | -1.30 | -4.94, 2.34 | 0.5 |
| Employment Status | | | | | | |
| Retired and not currently working (reference) | | | | | | |
| Not retired and not currently working | 0.087 | -3.20, 3.37 | >0.9 | -0.146 | -2.78, 2.48 | >0.9 |
| Employed <= 35h/wk | -0.366 | -3.81, 3.08 | 0.8 | -0.211 | -3.02, 2.60 | 0.9 |
| Employed > 35h/wk | -1.31 | -4.66, 2.03 | 0.4 | -0.353 | -3.16, 2.45 | 0.8 |
| Education (Less than HS) | -0.482 | -2.27, 1.31 | 0.6 | 0.155 | -1.14, 1.45 | 0.8 |
| Age at Immigration | | | | | | |
| >= 21 years (reference) | | | | | | |
| US Born | N/A | N/A | N/A | N/A | N/A | N/A |
| 0-5 years | 4.22 | -13.7, 22.1 | 0.6 | -8.80 | -14.9, -2.71 | 0.005 |
| 6-12 years | 0.795 | -1.95, 3.54 | 0.6 | 3.24 | -1.67, 8.16 | 0.2 |
| 13-20 years | 1.51 | -1.31, 4.33 | 0.3 | 1.36 | -0.211, 2.93 | 0.091 |
| Healthy Diet (Top 40th Percentile Diet Score) | -1.26 | -2.62, 0.104 | 0.072 | -0.127 | -1.33, 1.08 | 0.8 |
| Income | -1.47 | -2.80, -0.148 | 0.031 | -0.548 | -1.72, 0.622 | 0.4 |
| $PGS_{BMI}$ * Healthy Diet | -0.514 | -1.87, 0.838 | 0.5 | 0.351 | -0.849, 1.55 | 0.6 |
| $PGS_{BMI}$ * Age at immigration (Reference = >=21 years) | | | | | | |
| $PGS_{BMI}$ * US Born | -1.34 | -3.75,1.07 | 0.3 | 5.03 | -1.80,11.9 | 0.2 |
| $PGS_{BMI}$ * 0-5 years | 3.39 | -16.3,23.1 | 0.7 | N/A | N/A | N/A |
| $PGS_{BMI}$ * 6-12 years | 1.66 | -0.421,3.74 | 0.12 | 0.507 | -8.21,9.22 | >0.9 |
| $PGS_{BMI}$ * 13-20 years | -0.214 | -3.06,2.63 | 0.9 | -0.666 | -2.25,0.914 | 0.4 |

Supplemental Table 4. Complete Population Characteristics of the HCHS/SOL Analytic Subsample Overall (unweighted n=7,282) and Stratified by Background Group

| Characteristic | Overall[1] | Central American | Cuban | Dominican | Mexican | Puerto Rican | South American | p-value |
|---|---|---|---|---|---|---|---|---|
| Total | 7282 | 846 | 1331 | 659 | 2701 | 1211 | 534 | -- |
| Weighted | 7075 | 602 | 1717 | 636 | 2560 | 1157 | 403 | -- |
| AGE (years) | 43 (33, 54) | 40 (32, 51) | 48 (39, 59) | 43 (33, 53) | 40 (31, 51) | 45 (34, 56) | 44 (36, 53) | <0.001 |
| STUDY CENTER | | | | | | | | <0.001 |
| Brooklyn | 1,965 (28%) | 81 (13%) | 117 (6.8%) | 594 (93%) | 252 (9.8%) | 825 (71%) | 96 (24%) | |
| Chicago | 1,033 (15%) | 111 (19%) | 21 (1.2%) | 6 (0.9%) | 613 (24%) | 219 (19%) | 63 (16%) | |
| Miami | 2,287 (32%) | 358 (59%) | 1,567 (91%) | 30 (4.7%) | 43 (1.7%) | 76 (6.6%) | 214 (53%) | |
| San Diego | 1,790 (25%) | 52 (8.6%) | 12 (0.7%) | 6 (0.9%) | 1,653 (65%) | 38 (3.3%) | 30 (7.4%) | |
| US_BORN | 1,255 (18%) | 36 (5.9%) | 144 (8.4%) | 48 (7.5%) | 478 (19%) | 514 (44%) | 36 (9.0%) | <0.001 |
| YEARS LIVED IN THE US | 19 (8, 30) | 14 (7, 22) | 11 (4, 26) | 17 (10, 25) | 19 (10, 30) | 37 (26, 47) | 12 (8, 23) | <0.001 |
| AGE AT MIGRATION | | | | | | | | <0.001 |
| Born in the US | 1,255 (18%) | 36 (5.9%) | 144 (8.4%) | 48 (7.5%) | 478 (19%) | 514 (44%) | 36 (9.0%) | |
| 0 - <6 yrs | 337 (4.8%) | 8 (1.3%) | 35 (2.0%) | 19 (3.0%) | 114 (4.5%) | 153 (13%) | 8 (1.9%) | |

| | | | | | | | | |
|---|---|---|---|---|---|---|---|---|
| 6 - 12 yrs | 326 (4.6%) | 25 (4.1%) | 67 (3.9%) | 33 (5.1%) | 111 (4.3%) | 79 (6.8%) | 12 (3.0%) | |
| 13 - 20 yrs | 1,173 (17%) | 121 (20%) | 141 (8.2%) | 119 (19%) | 582 (23%) | 167 (14%) | 42 (10%) | |
| >=21 yrs | 3,985 (56%) | 413 (69%) | 1,330 (77%) | 417 (66%) | 1,275 (50%) | 245 (21%) | 306 (76%) | |
| IMMIGRANT GENERATION | | | | | | | | <0.001 |
| 1st Generation | 5,714 (81%) | 567 (94%) | 1,570 (91%) | 587 (92%) | 2,012 (79%) | 612 (53%) | 367 (91%) | |
| 2nd Generation or beyond | 1,361 (19%) | 36 (5.9%) | 147 (8.6%) | 48 (7.6%) | 548 (21%) | 546 (47%) | 36 (9.0%) | |
| SASH LANGUAGE SCORE | 1.50 (1.00, 2.67) | 1.33 (1.00, 2.00) | 1.17 (1.00, 1.83) | 1.50 (1.00, 2.17) | 1.67 (1.00, 2.67) | 3.17 (2.00, 4.00) | 1.50 (1.17, 2.17) | <0.001 |
| SASH SOCIAL SCORE | 2.25 (1.75, 2.75) | 2.00 (1.50, 2.50) | 2.00 (1.50, 2.50) | 2.25 (2.00, 2.75) | 2.25 (1.75, 2.75) | 2.50 (2.25, 3.00) | 2.25 (1.75, 2.75) | <0.001 |
| EDUCATION | | | | | | | | <0.001 |
| Less than HS | 2,160 (31%) | 227 (38%) | 339 (20%) | 230 (36%) | 903 (35%) | 390 (34%) | 72 (18%) | |
| >= HS | 4,916 (69%) | 375 (62%) | 1,379 (80%) | 405 (64%) | 1,657 (65%) | 768 (66%) | 332 (82%) | |
| Married, living with spouse | 4,047 (57%) | 361 (60%) | 948 (55%) | 290 (46%) | 1,778 (69%) | 437 (38%) | 233 (58%) | <0.001 |
| INCOME | | | | | | | | <0.001 |
| <$30,000 USD | 4,626 (65%) | 454 (75%) | 1,235 (72%) | 462 (73%) | 1,481 (58%) | 739 (64%) | 257 (64%) | |

| | | | | | | | | |
|---|---|---|---|---|---|---|---|---|
| >= $30,000 USD | 2,450 (35%) | 148 (25%) | 483 (28%) | 174 (27%) | 1,079 (42%) | 419 (36%) | 147 (36%) | |
| EMPLOYMENT STATUS | | | | | | | | <0.001 |
| Retired and not currently working | 649 (9.2%) | 25 (4.1%) | 202 (12%) | 65 (10%) | 122 (4.8%) | 209 (18%) | 27 (6.6%) | |
| Not retired and not currently working | 2,581 (36%) | 193 (32%) | 735 (43%) | 232 (37%) | 883 (34%) | 422 (36%) | 116 (29%) | |
| Employed <= 35h/wk | 1,190 (17%) | 141 (23%) | 194 (11%) | 115 (18%) | 501 (20%) | 152 (13%) | 86 (21%) | |
| Employed > 35h/wk | 2,656 (38%) | 244 (40%) | 586 (34%) | 224 (35%) | 1,054 (41%) | 374 (32%) | 174 (43%) | |
| Ethnic Identity Summary Score | | | | | | | | <0.001 |
| 1 | 36 (0.5%) | 3 (0.5%) | 6 (0.4%) | 1 (0.2%) | 15 (0.6%) | 9 (0.8%) | 2 (0.4%) | |
| 1.5 | 34 (0.5%) | 3 (0.5%) | 10 (0.6%) | 0 (0%) | 14 (0.5%) | 5 (0.4%) | 2 (0.4%) | |
| 2 | 252 (3.6%) | 30 (4.9%) | 38 (2.2%) | 9 (1.4%) | 100 (3.9%) | 61 (5.2%) | 15 (3.7%) | |
| 2.5 | 834 (12%) | 40 (6.7%) | 110 (6.4%) | 83 (13%) | 361 (14%) | 188 (16%) | 51 (13%) | |
| 3 | 3,313 (47%) | 273 (45%) | 856 (50%) | 285 (45%) | 1,181 (46%) | 555 (48%) | 162 (40%) | |
| 3.5 | 1,034 (15%) | 80 (13%) | 218 (13%) | 111 (17%) | 409 (16%) | 148 (13%) | 67 (17%) | |
| 4 | 1,573 (22%) | 172 (29%) | 478 (28%) | 146 (23%) | 479 (19%) | 192 (17%) | 106 (26%) | |
| CES-D 10 Item Summary Score | 6.0 (3.0, 10.0) | 5.0 (2.0, 9.0) | 5.0 (2.0, 11.0) | 6.0 (2.0, 11.0) | 5.0 (3.0, 9.0) | 7.0 (4.0, 11.0) | 6.0 (2.0, 10.0) | <0.001 |

| | | | | | | | | |
|---|---|---|---|---|---|---|---|---|
| Sweetened Beverage Consumption (Servings/day) | 1.58 (0.97, 2.34) | 1.73 (1.06, 2.61) | 1.48 (0.95, 2.11) | 1.31 (0.74, 1.85) | 1.59 (0.97, 2.38) | 1.77 (1.12, 2.66) | 1.64 (1.02, 2.37) | <0.001 |
| Meets 2008 Physical Activity Guidelines | 4,703 (66%) | 421 (70%) | 970 (56%) | 424 (67%) | 1,807 (71%) | 795 (69%) | 287 (71%) | <0.001 |
| JAMA Healthy Diet Score | | | | | | | | <0.001 |
| <=60th sex-specific percentile | 3,683 (52%) | 339 (56%) | 998 (58%) | 493 (78%) | 733 (29%) | 927 (80%) | 193 (48%) | |
| >60th sex-specific percentile | 3,393 (48%) | 264 (44%) | 719 (42%) | 143 (22%) | 1,827 (71%) | 231 (20%) | 210 (52%) | |
| Sleep Duration (h/day) | 7.93 (7.00, 8.71) | 7.79 (7.00, 8.57) | 8.00 (7.21, 8.93) | 7.86 (7.00, 8.93) | 8.00 (7.29, 8.71) | 7.64 (6.71, 8.79) | 7.71 (6.79, 8.50) | <0.001 |
| CIGARETTE USE | | | | | | | | <0.001 |
| Never | 4,223 (60%) | 390 (65%) | 938 (55%) | 501 (79%) | 1,586 (62%) | 570 (49%) | 236 (59%) | |
| Former | 1,365 (19%) | 119 (20%) | 342 (20%) | 80 (13%) | 524 (20%) | 191 (17%) | 109 (27%) | |
| Current | 1,489 (21%) | 93 (15%) | 437 (25%) | 55 (8.6%) | 450 (18%) | 396 (34%) | 58 (14%) | |
| ALCOHOL USE LEVEL | | | | | | | | 0.004 |
| Non-drinker | 3,382 (48%) | 336 (56%) | 857 (50%) | 290 (46%) | 1,157 (45%) | 567 (49%) | 175 (43%) | |

| | | | | | | | | |
|---|---|---|---|---|---|---|---|---|
| Low-risk drinker | 3,252 (46%) | 228 (38%) | 745 (43%) | 312 (49%) | 1,222 (48%) | 527 (46%) | 218 (54%) | |
| At-risk drinker | 442 (6.2%) | 38 (6.3%) | 115 (6.7%) | 34 (5.4%) | 181 (7.1%) | 63 (5.5%) | 10 (2.6%) | |
| Diabetes History | 1,154 (16%) | 87 (14%) | 298 (17%) | 96 (15%) | 417 (16%) | 216 (19%) | 41 (10%) | 0.025 |
| Prevalent CVD | 462 (6.5%) | 37 (6.1%) | 151 (8.8%) | 40 (6.2%) | 123 (4.8%) | 92 (7.9%) | 20 (4.8%) | 0.055 |
| BMI | 28.9 (25.9, 32.7) | 28.8 (26.2, 32.4) | 29.0 (25.7, 32.6) | 29.3 (26.0, 32.7) | 28.7 (26.1, 32.3) | 29.6 (26.1, 34.3) | 27.9 (24.9, 30.8) | <0.001 |
| Waist-to-Hip Ratio | 0.93 (0.88, 0.97) | 0.93 (0.88, 0.96) | 0.92 (0.87, 0.97) | 0.91 (0.85, 0.96) | 0.94 (0.89, 0.97) | 0.92 (0.87, 0.97) | 0.91 (0.86, 0.96) | <0.001 |
| Obesity | 2,948 (42%) | 238 (39%) | 716 (42%) | 268 (42%) | 1,054 (41%) | 550 (48%) | 123 (31%) | 0.002 |
| BMI PRS | 0.01 (-0.65, 0.67) | 0.07 (-0.54, 0.67) | -0.14 (-0.88,0.55) | 0.09 (-0.55, 0.69) | 0.07 (-0.61, 0.72) | -0.03 (-0.69, 0.64) | -0.02 (-0.64, 0.62) | <0.001 |

[1] ***Median (IQR); n (%)***

Supplemental Table 5. Complete Amerindigenous Ancestry Univariate Associations with Environmental Variables

| | Overall (n=7075) | | | Central American (n=602) | | | Cuban American (n=1717) | | | Dominican (n=635) | | | Mexican (n=2560) | | | Puerto Rican (n=1157) | | | South American (n=403) | | |
|---|---|---|---|---|---|---|---|---|---|---|---|---|---|---|---|---|---|---|---|---|---|
| **Characteristic** | **Beta** | **95% CI** | **p-value** | **Beta** | **95% CI** | **p-value** | **Beta** | **95% CI** | **p-value** | **Beta** | **95% CI** | **p-value** | **Beta** | **95% CI** | **p-value** | **Beta** | **95% CI** | **p-value** | **Beta** | **95% CI** | **p-value** |
| **AGE** | -0.003 | -0.004, -0.003 | **<0.001** | -0.001 | -0.002, 0.000 | **0.007** | -0.001 | -0.001, 0.000 | **<0.001** | 0.000 | 0.000, 0.000 | 0.44 | -0.003 | -0.003, -0.002 | **<0.001** | 0.000 | 0.000, 0.000 | **0.024** | -0.002 | -0.004, 0.000 | **0.038** |
| **STUDY CENTER** | | | | | | | | | | | | | | | | | | | | | |
| Brooklyn | | | | | | | | | | | | | | | | | | | | | |
| Chicago | 0.300 | 0.286, 0.313 | **<0.001** | 0.009 | -0.032, 0.050 | 0.67 | 0.014 | -0.005, 0.033 | 0.15 | -0.007 | -0.020, 0.006 | 0.29 | -0.207 | -0.238, -0.177 | **<0.001** | 0.011 | 0.007, 0.016 | **<0.001** | 0.091 | 0.035, 0.146 | **0.001** |
| Miami | 0.017 | 0.004, 0.030 | **0.009** | -0.059 | -0.098, -0.021 | **0.003** | -0.002 | -0.012, 0.008 | 0.68 | -0.001 | -0.010, 0.008 | 0.84 | -0.333 | -0.399, -0.266 | **<0.001** | -0.002 | -0.011, 0.008 | 0.73 | -0.135 | -0.186, -0.084 | **<0.001** |
| San Diego | 0.248 | 0.235, 0.262 | **<0.001** | -0.041 | -0.095, 0.014 | 0.14 | 0.029 | 0.003, 0.056 | **0.027** | 0.008 | -0.025, 0.040 | 0.64 | -0.351 | -0.381, -0.321 | **<0.001** | 0.007 | -0.008, 0.022 | 0.38 | -0.238 | -0.337, -0.139 | **<0.001** |
| **US BORN** | -0.083 | -0.099, -0.067 | **<0.001** | -0.100 | -0.156, -0.044 | **<0.001** | 0.005 | -0.005, 0.015 | 0.30 | -0.006 | -0.015, 0.002 | 0.13 | -0.091 | -0.111, -0.072 | **<0.001** | -0.005 | -0.008, -0.001 | **0.024** | -0.162 | -0.257, -0.068 | **<0.001** |
| **Years Lived in the US** | -0.002 | -0.002, -0.002 | **<0.001** | -0.001 | -0.002, 0.000 | **0.029** | 0.000 | -0.001, 0.000 | **<0.001** | 0.0 | 0.000, 0.000 | 0.055 | -0.0 | -0.003, -0.002 | **<0.001** | 0.0 | 0.000, 0.000 | 0.27 | -0.002 | -0.004, 0.000 | **0.038** |

| | | | | | | | | | | | | | | | | | | | | | |
|---|---|---|---|---|---|---|---|---|---|---|---|---|---|---|---|---|---|---|---|---|---|
| | | | | | | | | | | 00 | | | 03 | | | 00 | | | | | |
| **>=21 years** | REF | REF | REF | REF | REF | REF | REF | REF | REF | REF | REF | REF | REF | REF | REF | REF | REF | REF | REF | REF | REF |
| **US BORN** | -0.079 | -0.095, -0.062 | **<0.001** | -0.098 | -0.154, -0.041 | **<0.001** | 0.005 | -0.005,0.014 | 0.37 | -0.006 | -0.015, 0.002 | 0.13 | -0.094 | -0.114, -0.074 | **<0.001** | -0.003 | -0.008, 0.002 | 0.26 | -0.153 | -0.247, -0.060 | **0.001** |
| **0-5 years** | -0.082 | -0.112, -0.052 | **<0.001** | -0.003 | -0.109, 0.102 | 0.95 | -0.006 | -0.024,0.012 | 0.51 | -0.002 | -0.013, 0.009 | 0.70 | -0.054 | -0.093, -0.0015 | **0.006** | 0.002 | -0.005, 0.009 | 0.64 | -0.167 | -0.400, 0.066 | 0.16 |
| **6-12 years** | -0.052 | -0.079, -0.025 | **<0.001** | -0.017 | -0.084, 0.051 | 0.63 | -0.017 | -0.031, -0.003 | **0.017** | 0.004 | -0.005, 0.013 | 0.39 | -0.020 | -0.054, 0.014 | 0.24 | 0.006 | -0.002, 0.013 | 0.13 | -0.065 | -0.201, 0.071 | 0.35 |
| **13-20 years** | 0.059 | 0.043, 0.074 | **<0.001** | 0.021 | -0.008, 0.050 | 0.15 | 0.000 | -0.010, 0.010 | >0.99 | -0.001 | -0.006, 0.004 | 0.68 | 0.002 | -0.015, 0.019 | 0.81 | 0.002 | -0.004, 0.008 | 0.48 | 0.110 | 0.044,0.176 | **0.001** |
| **Immigrant Generation** | -0.075 | -0.091, -0.060 | **<0.001** | -0.100 | -0.156, -0.044 | **<0.001** | 0.005 | -0.005, 0.014 | 0.32 | -0.008 | -0.016, 0.001 | 0.067 | -0.098 | -0.115, -0.080 | **<0.001** | -0.005 | -0.009, -0.001 | **0.018** | -0.162 | -0.257, -0.068 | **<0.001** |
| **SASH LANG** | -0.033 | -0.038, -0.027 | **<0.001** | -0.014 | -0.028, 0.000 | **0.048** | -0.002 | -0.004, 0.001 | 0.30 | -0.002 | -0.005, 0.000 | **0.047** | -0.037 | -0.044, -0.031 | **<0.001** | -0.001 | -0.002, 0.001 | 0.30 | -0.043 | -0.070, -0.017 | **0.002** |
| **SASH SOC** | -0.035 | -0.044, -0.025 | **<0.001** | -0.005 | -0.022, 0.013 | 0.59 | 0.000 | -0.005, 0.004 | 0.97 | -0.001 | -0.005, 0.002 | 0.40 | -0.029 | -0.040, -0.018 | **<0.001** | -0.002 | -0.006, 0.002 | 0.27 | -0.007 | -0.043, 0.029 | 0.71 |

| | | | | | | | | | | | | | | | | | | | | | |
|---|---|---|---|---|---|---|---|---|---|---|---|---|---|---|---|---|---|---|---|---|---|
| **EDUCATION** | -0.078 | -0.090, -0.066 | **<0.001** | -0.047 | -0.068, -0.026 | **<0.001** | 0.000 | -0.006, 0.006 | 0.97 | -0.005 | -0.009, -0.002 | **0.003** | -0.064 | -0.077, -0.051 | **<0.001** | -0.004 | -0.008, 0.000 | 0.078 | -0.158 | -0.206, -0.109 | **<0.001** |
| **Married, living with spouse** | 0.077 | 0.066, 0.088 | **<0.001** | 0.007 | -0.014, 0.028 | 0.51 | 0.001 | -0.004, 0.006 | 0.80 | 0.000 | -0.003, 0.004 | 0.86 | 0.014 | 0.000, 0.029 | 0.057 | 0.005 | 0.001, 0.009 | **0.018** | 0.011 | -0.031, 0.052 | 0.62 |
| **INCOME** | 0.004 | -0.008, 0.016 | 0.55 | -0.017 | -0.041, 0.007 | 0.17 | 0.001 | -0.005, 0.006 | 0.79 | -0.001 | -0.005, 0.003 | 0.60 | -0.060 | -0.074, -0.046 | **<0.001** | 0.001 | -0.003, 0.006 | 0.49 | -0.015 | -0.060, 0.029 | 0.50 |
| **EMPLOYMENT STATUS** | | | | | | | | | | | | | | | | | | | | | |
| Retired and not currently working | REF | REF | REF | REF | REF | REF | REF | REF | REF | REF | REF | REF | REF | REF | REF | REF | REF | REF | REF | REF | REF |
| Not retired and not currently working | 0.062 | 0.042, 0.082 | **<0.001** | -0.018 | -0.066, 0.031 | 0.47 | 0.011 | 0.002, 0.019 | **0.019** | 0.004 | -0.002, 0.010 | 0.16 | 0.034 | 0.005, 0.062 | **0.020** | 0.001 | -0.004, 0.006 | 0.65 | 0.003 | -0.081, 0.087 | 0.95 |
| Employed <= 35h/wk | 0.133 | 0.110, 0.155 | **<0.001** | 0.000 | -0.050, 0.051 | 0.99 | 0.010 | -0.001, 0.020 | 0.084 | 0.005 | -0.001, 0.012 | 0.12 | 0.044 | 0.014, 0.075 | **0.004** | -0.006 | -0.014, 0.001 | 0.11 | 0.001 | -0.085, 0.087 | 0.98 |
| Employed > 35h/wk | 0.107 | 0.087, 0.126 | **<0.001** | -0.002 | -0.050, 0.046 | 0.95 | 0.012 | 0.002, 0.021 | **0.013** | 0.005 | -0.001, 0.011 | 0.090 | 0.046 | 0.018, 0.074 | **0.002** | 0.002 | -0.004, 0.007 | 0.55 | 0.018 | -0.063, 0.100 | 0.66 |
| **Ethnic Identity Summary Score** | 0.005 | -0.005, 0.015 | 0.35 | 0.000 | -0.018, 0.018 | 0.98 | 0.002 | -0.003, 0.006 | 0.50 | 0.000 | -0.004, 0.003 | 0.89 | 0.017 | 0.006, 0.029 | **0.004** | -0.001 | -0.004, 0.003 | 0.69 | 0.014 | -0.021, 0.049 | 0.42 |

| | | | | | | | | | | | | | | | | | | | | | |
|---|---|---|---|---|---|---|---|---|---|---|---|---|---|---|---|---|---|---|---|---|---|
| **CES-D 10 Item Score** | -0.003 | -0.004, -0.002 | **<0.001** | -0.001 | -0.003, 0.000 | 0.12 | 0.000 | 0.000, 0.000 | 0.72 | 0.000 | 0.000, 0.000 | 0.81 | 0.000 | -0.002, 0.001 | 0.53 | 0.000 | 0.000, 0.000 | 0.53 | 0.002 | -0.002, 0.006 | 0.27 |
| **Sweetened Beverage Consumption (Servings/day)** | 0.016 | 0.011, 0.022 | **<0.001** | 0.000 | -0.010, 0.010 | >0.99 | 0.003 | 0.000, 0.006 | **0.038** | -0.001 | -0.003, 0.001 | 0.38 | 0.018 | 0.011, 0.024 | **<0.001** | 0.001 | -0.001, 0.003 | 0.22 | 0.020 | 0.000, 0.040 | 0.052 |
| **Meets 2008 Physical Activity Guidelines** | 0.037 | 0.025, 0.049 | **<0.001** | 0.000 | -0.022, 0.022 | >0.99 | 0.000 | -0.005, 0.005 | 0.96 | -0.001 | -0.005, 0.002 | 0.44 | 0.008 | -0.007, 0.022 | 0.29 | -0.001 | -0.005, 0.003 | 0.64 | 0.017 | -0.027, 0.061 | 0.45 |
| **Top 40th Diet Score** | 0.135 | 0.124, 0.146 | **<0.001** | -0.002 | -0.023, 0.018 | 0.82 | -0.002 | -0.007, 0.003 | 0.36 | 0.000 | -0.004, 0.004 | 0.89 | -0.030 | -0.046, -0.014 | **<0.001** | 0.001 | -0.004, 0.005 | 0.82 | -0.017 | -0.058, 0.024 | 0.42 |
| **SLPDUR** | -0.001 | -0.005, 0.003 | 0.59 | -0.011 | -0.019, -0.003 | **0.005** | 0.000 | -0.002, 0.002 | 0.82 | 0.001 | 0.000, 0.002 | 0.16 | -0.002 | -0.007, 0.004 | 0.53 | 0.000 | -0.001, 0.001 | 0.85 | -0.034 | -0.050, -0.019 | **<0.001** |
| **CIGARETTE_USE** | | | | | | | | | | | | | | | | | | | | | |
| Never | REF | REF | REF | REF | REF | REF | REF | REF | REF | REF | REF | REF | REF | REF | REF | REF | REF | REF | REF | REF | REF |
| Former | -0.025 | -0.039, -0.011 | **<0.001** | -0.048 | -0.073, -0.023 | **<0.001** | -0.005 | -0.011, 0.002 | 0.15 | -0.004 | -0.009, 0.001 | 0.13 | -0.037 | -0.054, -0.021 | **<0.001** | -0.001 | -0.006, 0.004 | 0.74 | -0.057 | -0.104, -0.009 | **0.020** |
| Current | -0.090 | -0.104, -0.075 | **<0.001** | -0.020 | -0.052, 0.011 | 0.20 | -0.006 | -0.012, 0.000 | 0.051 | -0.002 | -0.008, 0.004 | 0.59 | -0.048 | -0.067, -0.029 | **<0.001** | -0.002 | -0.006, 0.003 | 0.49 | -0.135 | -0.196, -0.074 | **<0.001** |

| | | | | | | | | | | | | | | | | | | | | | |
|---|---|---|---|---|---|---|---|---|---|---|---|---|---|---|---|---|---|---|---|---|---|
| **Alcohol Use Level** | | | | | | | | | | | | | | | | | | | | | |
| Non-drinker | REF | REF | REF | REF | REF | REF | REF | REF | REF | REF | REF | REF | REF | REF | REF | REF | REF | REF | REF | REF | REF |
| Low-risk drinker | -0.013 | -0.025, -0.002 | **0.026** | -0.011 | -0.033, 0.011 | 0.33 | 0.002 | -0.003, 0.007 | 0.51 | -0.001 | -0.004, 0.003 | 0.67 | -0.023 | -0.037, -0.009 | **0.001** | 0.000 | -0.004, 0.003 | 0.81 | -0.059 | -0.100, -0.018 | **0.005** |
| At-risk drinker | -0.031 | -0.057, -0.005 | **0.020** | -0.014 | -0.062, 0.033 | 0.55 | 0.006 | -0.004, 0.017 | 0.25 | 0.001 | -0.008, 0.009 | 0.89 | -0.041 | -0.071, -0.010 | **0.009** | 0.007 | -0.002, 0.016 | 0.11 | -0.164 | -0.308, -0.020 | **0.026** |
| **Diabetes History** | 0.000 | -0.014, 0.014 | 0.97 | -0.014 | -0.041, 0.013 | 0.30 | -0.006 | -0.012, 0.001 | 0.076 | 0.000 | -0.005, 0.004 | 0.86 | 0.025 | 0.008, 0.042 | **0.003** | 0.003 | -0.001, 0.007 | 0.19 | 0.016 | -0.044, 0.076 | 0.61 |
| **Prevalent CVD** | -0.054 | -0.076, -0.031 | **<0.001** | 0.012 | -0.036, 0.060 | 0.62 | -0.005 | -0.014, 0.004 | 0.31 | -0.006 | -0.013, 0.000 | 0.070 | 0.001 | -0.030, 0.031 | 0.95 | 0.001 | -0.006, 0.008 | 0.76 | -0.010 | -0.098, 0.077 | 0.82 |
| **BMI** | -0.001 | -0.002, 0.000 | **0.044** | -0.002 | -0.004, 0.000 | 0.076 | 0.000 | 0.000, 0.001 | 0.68 | 0.000 | 0.000, 0.000 | 0.84 | 0.000 | -0.001, 0.001 | 0.97 | 0.000 | 0.000, 0.000 | 0.20 | 0.000 | -0.004, 0.004 | 0.88 |
| **Waist-hip ratio** | 0.204 | 0.127, 0.281 | **<0.001** | -0.028 | -0.171, 0.115 | 0.70 | -0.048 | -0.079, -0.017 | **0.003** | 0.007 | -0.016, 0.029 | 0.56 | 0.215 | 0.114, 0.316 | **<0.001** | 0.001 | -0.025, 0.027 | 0.93 | 0.426 | 0.158, 0.694 | **0.002** |
| **Obesity** | -0.004 | -0.015, 0.007 | 0.49 | -0.017 | -0.038, 0.004 | 0.10 | 0.001 | -0.004, 0.007 | 0.56 | -0.004 | -0.007, 0.000 | 0.056 | 0.010 | -0.004, 0.024 | 0.15 | -0.002 | -0.005, 0.002 | 0.41 | 0.015 | -0.028, 0.058 | 0.50 |

| | | | | | | | | | | | | | | | | | | | | | |
|---|---|---|---|---|---|---|---|---|---|---|---|---|---|---|---|---|---|---|---|---|---|
| **BMI PGS** | 0.030 | 0.024, 0.036 | **<0.001** | 0.018 | 0.007, 0.029 | **0.001** | 0.005 | 0.002, 0.007 | **<0.001** | -0.003 | -0.004, -0.001 | **0.013** | 0.040 | 0.033, 0.047 | **<0.001** | -0.001 | -0.003, 0.001 | 0.39 | 0.036 | 0.014, 0.057 | **0.001** |

Supplementary Figure 1. Inclusion/Exclusion Criteria Flowchart

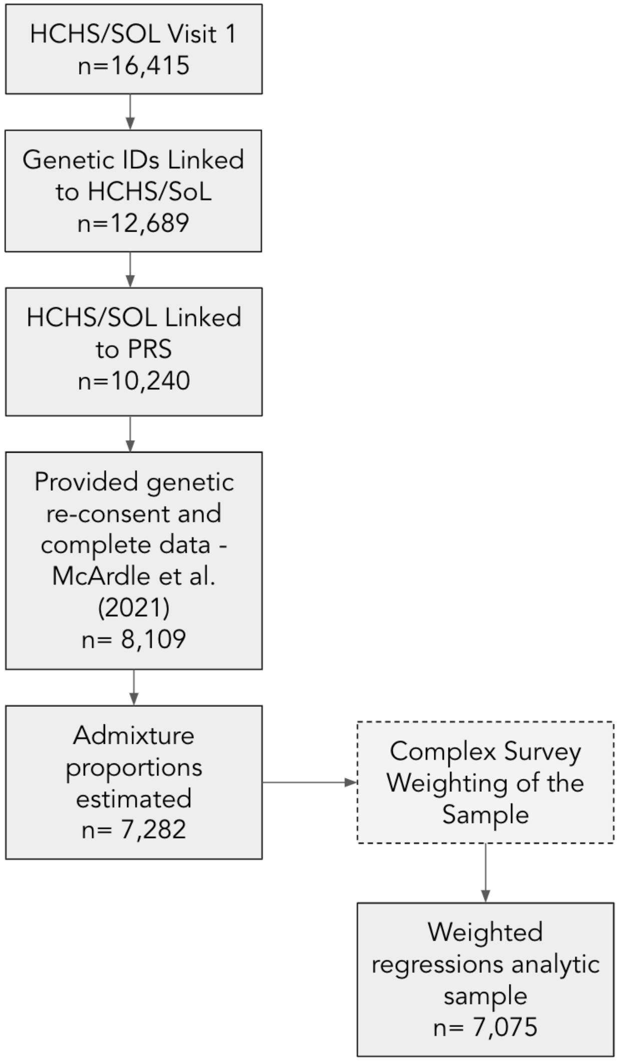

### Supplementary Figure 2. Distributions of Selected Variables Aggregated and by Background Group

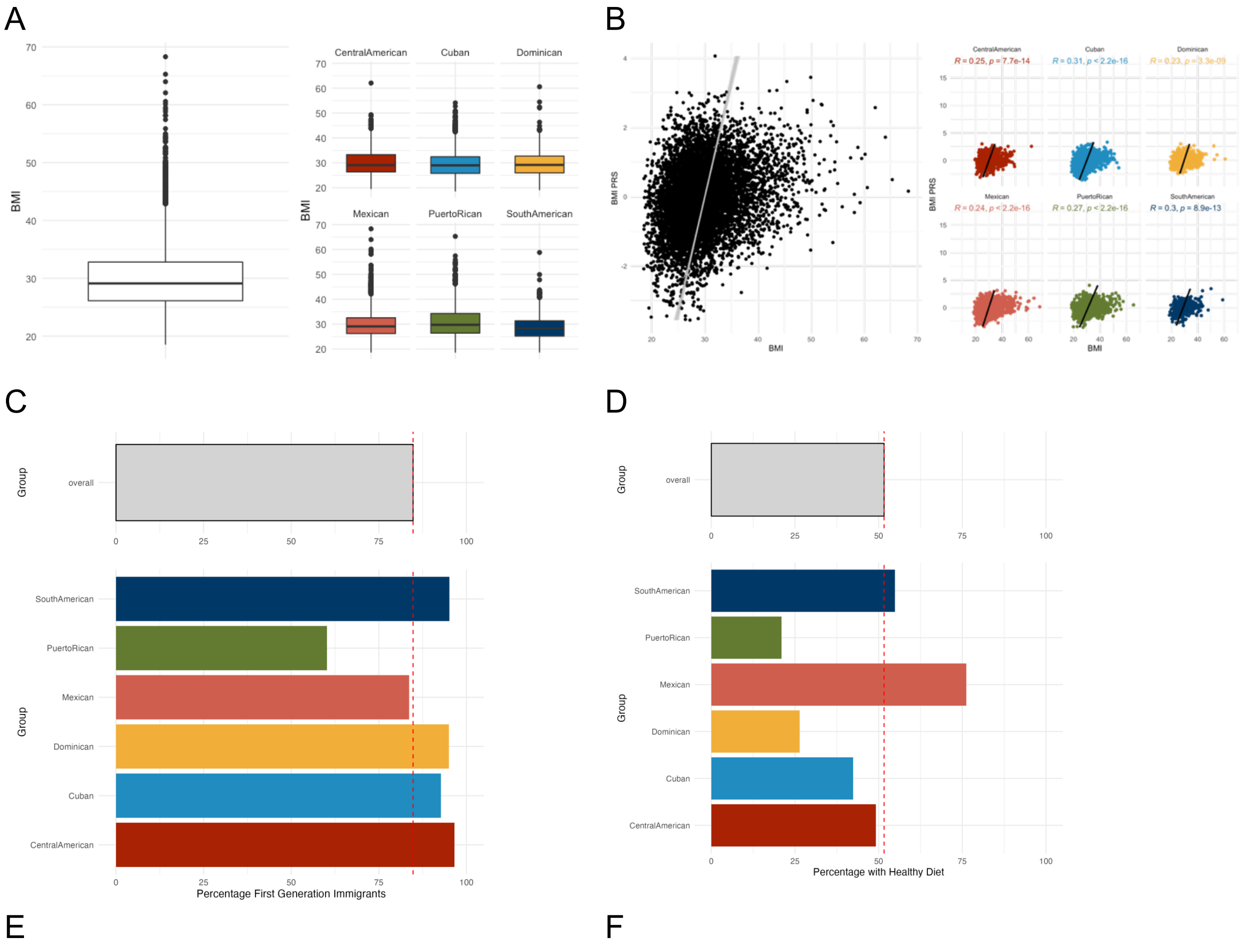

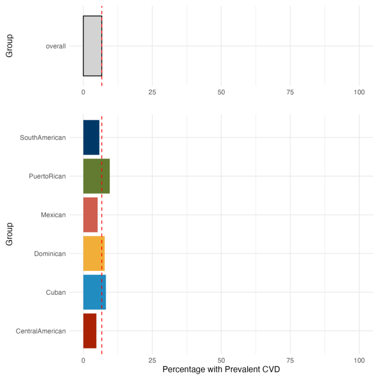


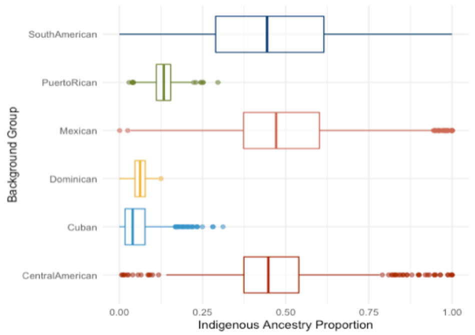


*Graphical depictions of distributions or proportions of A) BMI, B) PGS$_{BMI}$ vs. BMI, C) immigrant generation (1st generation or 2nd generation and beyond), D) JAMA Healthy Diet score (>=60th percentile score indicates healthy diet), and E) prevalent cardiovascular disease in the complete sample (top, light gray bars) and stratified by self-identified background group identity (colored panels). F) Distribution of AME ancestry proportion by self-identified Hispanic/Latino background.*

Supplementary Figure 3. PRS-Age at Immigration Interactions by AME Ancestry Tertile and Background Group Identity

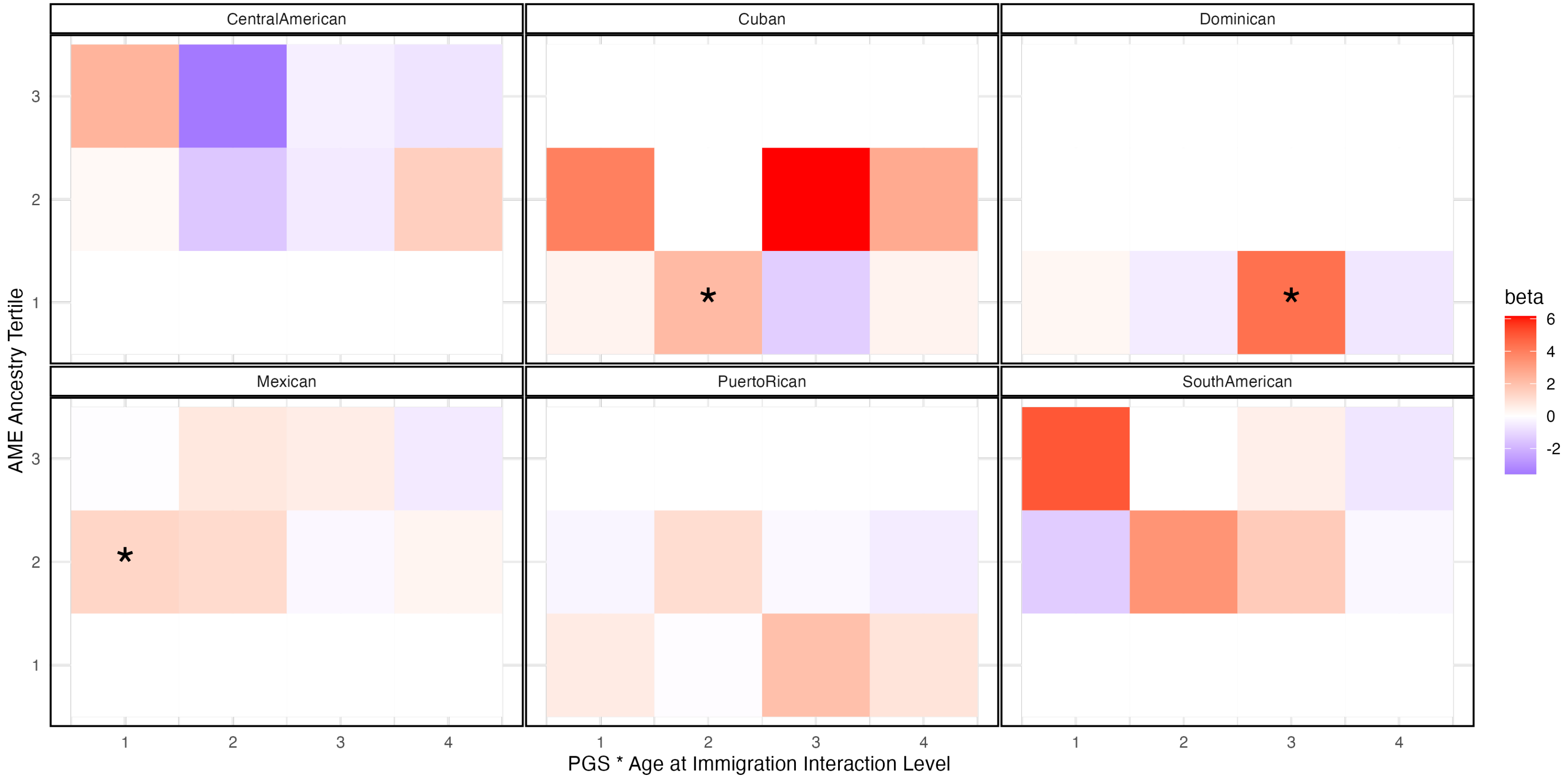


*Unshaded tiles indicate subgroups for which there was no sufficient sample size to observe PGS-Age at immigration interactions. PGS - Age at Immigration Interaction levels are as follows: (1) comparing those US-born to those who were ≥21 years at immigration, (2) comparing those 0-5 years at immigration to those ≥21 years at immigration, (3) comparing those 6-12 years at*

*immigration to those ≥21 years at immigration, and (4) comparing those 13-20 years at immigration to those who were ≥21 years at immigration. Significant contrasts are depicted with an asterisk (*).*
